# A comparison of Markov and mechanistic models for STH prevalence projections in the context of survey design

**DOI:** 10.1101/2023.10.02.23296429

**Authors:** Max T. Eyre, Caroline A. Bulstra, Olatunji Johnson, Sake J. de Vlas, Peter J. Diggle, Claudio Fronterrè, Luc E. Coffeng

## Abstract

Globally, there are over one billion people infected with soil-transmitted helminths (STHs), mostly living in marginalised settings with inadequate sanitation in sub-Saharan Africa and Southeast Asia. The WHO recommends an integrated approach to STH morbidity control through improved access to sanitation and hygiene education, and the delivery of preventive chemotherapy (PC) to school age children delivered through schools. Progress of STH control programmes is currently estimated using a baseline (pre-PC) school-based prevalence survey and then monitored using periodical school-based prevalence surveys, known as Impact Assessment Surveys (IAS). We investigated whether integrating geostatistical methods with a Markov model or a mechanistic transmission model for projecting prevalence forward in time from baseline can improve IAS design strategies. To do this, we applied these two methods to prevalence data collected in Kenya, before evaluating and comparing their performance in accurately informing optimal survey design for a range of IAS sampling designs. We found that although both approaches performed well, the mechanistic method more accurately projected prevalence over time and provided more accurate information for guiding survey design. Both methods performed less well in areas with persistent STH hotspots where prevalence did not reduce despite multiple rounds of PC. Our findings show that these methods can be useful tools for more efficient and accurate targeting of PC. The general framework built in this paper can also be used for projecting prevalence and informing survey design for other NTDs.

## Introduction

Soil-transmitted helminths (STHs) are parasitic intestinal nematodes that are transmitted between humans through contaminated soil and are composed of *Ascaris lumbricoides, Trichuris trichiura*, and hookworm *spp*. (*Ancylostoma duodenale* and *Necator americanus*). These four species are considered together because of their similar transmission dynamics, diagnosis, control, and prevention measures. It is common for a single individual, particularly children in impoverished settings, to be chronically infected with more than one species at the same time (1,2). Globally, there are over one billion people infected with STHs, one of the most common neglected tropical diseases (NTDs) worldwide. The majority of cases are found in marginalised settings with inadequate sanitation in sub-Saharan Africa and Southeast Asia and present a major public health burden globally (3).

The World Health Organization (WHO) recommends an integrated approach to STH morbidity control, through improved access to sanitation and hygiene education, and the school-based delivery of preventive chemotherapy (PC) with albendazole or mebendazole to school age children (SAC). It has set a target of reducing the prevalence of moderate and heavy intensity infections below 2% in SAC and pre-school age children (PSAC) by 2030 (4). Typically, STH prevalence is initially estimated using a baseline (pre-PC) school-based prevalence survey of SAC, conducted at a number of selected primary schools in endemic areas. STH control progress is then monitored using periodical school-based prevalence surveys, known as Impact Assessment Surveys (IAS). IAS are typically carried out after five years of PC and are used to estimate current STH prevalence at IU-level to inform decisions on the requirements for PC delivery with the aim of reaching elimination as a public health problem. When the prevalence of STH (any intensity) in the target population falls below 2%, the WHO recommends suspending PC (5).

In the context of limited financial resources in STH endemic countries and the high cost associated with conducting prevalence surveys, there is a need for careful design of surveys to accurately and efficiently measure prevalence burden and capture geographical variation in prevalence. Our previous work applying model-based geostatistical methods to this problem has demonstrated that they can significantly increase the precision of prevalence surveys relative to traditional survey design, thus reducing field-sampling effort while maintaining or improving precision (6,7). The aim of this study was to investigate whether integrating geostatistical methods with Markov or mechanistic models can accurately project prevalence forward in time and help improve IAS design. While this study focuses on STH impact surveys, the methodology and principles are applicable to post-baseline survey design for other NTDs.

## Methods

### Data

#### STH prevalence and PC coverage data

STH prevalence and PC coverage data were collected in 16 implementation units (IUs; districts, administrative level 2) in Kenya (see **Figure 1A**) to monitor the reduction in STH infection in response to annual PC for SAC during a national school-based deworming programme (NSBDP) between 2012 and 2017 (8). Estimated PC coverage in each round was based on pre-PC surveys carried out approximately one year after each previous PC round and 2-5 weeks before the start of the next PC round and were recorded for each IU (see **Supplementary Figure 1**). These data are publicly available via the Global Atlas of Helminth Infections (https://www.thiswormyworld.org/) and the ESPEN portal (https://espen.afro.who.int/).

**Figure 1.**
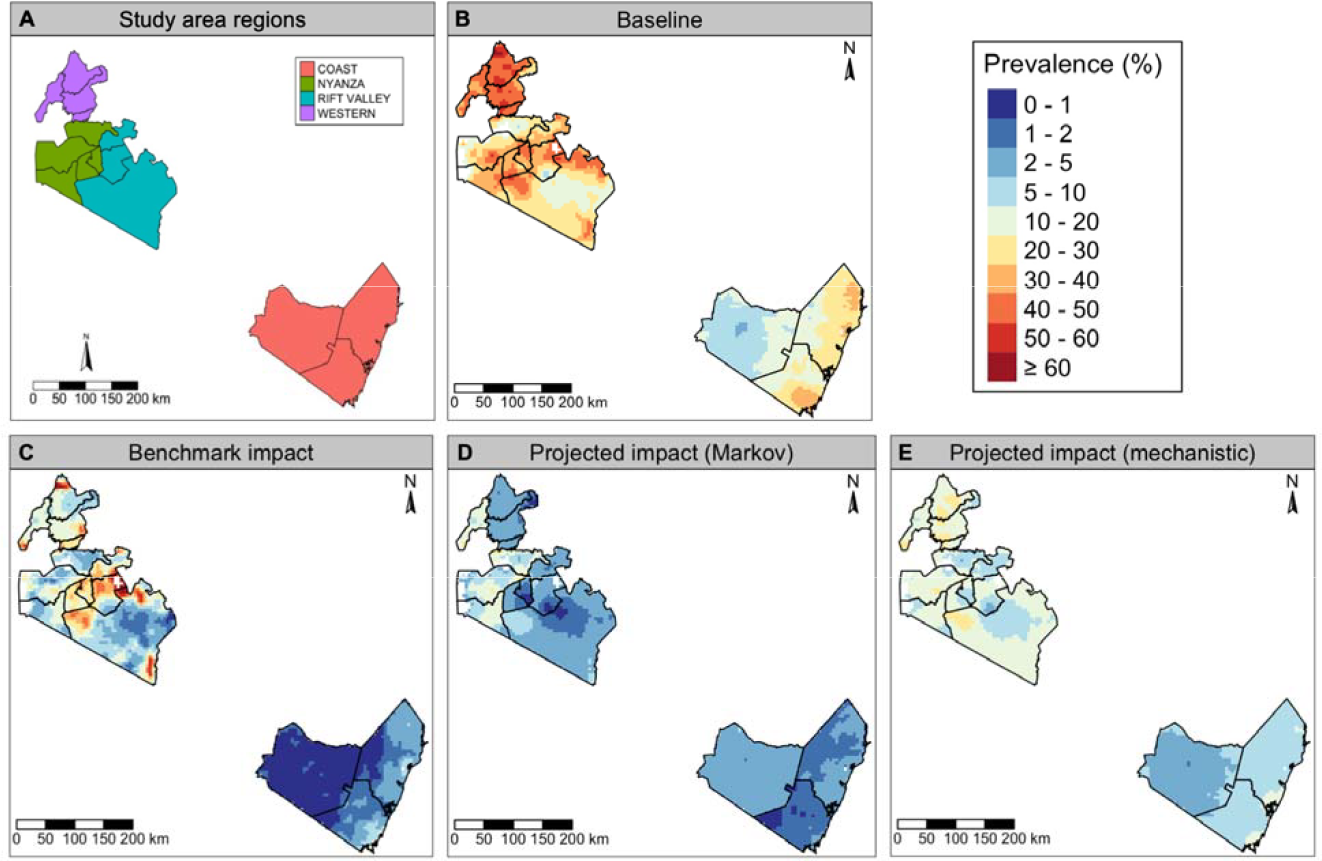
Panel A: Study area in Southwest Kenya, consisting of 16 implementation units (boundaries indicated by black lines) within four regions: Coast, Nyanza, Rift Valley and Western; Panels B-C: STH prevalence in SAC as predicted by the geostatistical model fit to baseline and impact prevalence survey data (the prediction surface at impact is used as our benchmark); Panels D-E: STH prevalence projected at impact using the Markov and mechanistic approaches.

The NSBDP study design (described in more detail previously (8)) consisted of repeat cross-sectional surveys in a representative, population-stratified random sample of 172 schools across the 16 IUs at three time points over a five-year period: baseline (pre-PC, 2012), midterm (after two rounds of PC, 2015) and impact (after four rounds of PC, 2017). During each survey, stool samples were collected from a randomly selected sample of approximately 100 school children at each school and tested for the presence of each STH species using the Kato-Katz thick smear technique.

#### Environmental and demographic data

Environmental data for the study area were available from MODIS (9,10) and consisted of rasters at 5 km resolution for the following variables: elevation, Enhanced Vegetation Index (EVI), mean daytime land surface temperature (LST), mean night-time LST, Normalized Difference Vegetation Index (NDVI), soil acidity and soil moisture. Population density data at 1km resolution was used from WorldPop (11).

### Overview of study analysis steps

This study consisted of three main steps (see **Supplementary Figure 2** for a diagram):

**Figure 2.**
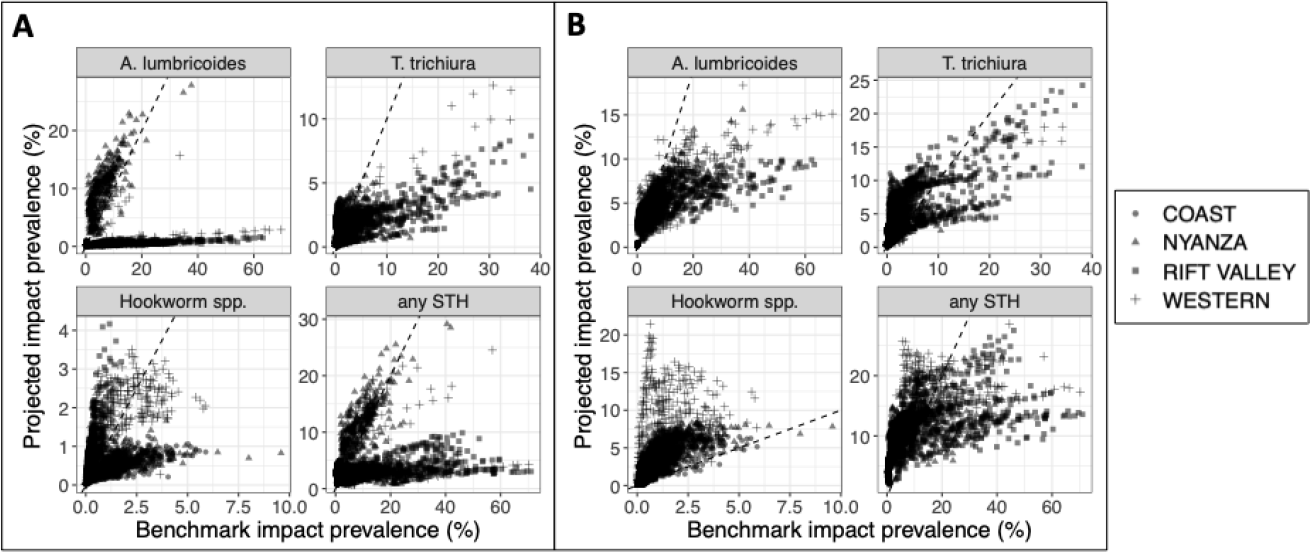
Prevalence predictions at impact projected by the Markov (Panel A) and mechanistic (Panel B) models compared to the benchmark values predicted by the geostatistical model fit to impact survey data, shown for each STH species with geographic region depicted by point shape (each point represents a pixel within the study area).

#### Step 1. Geostatistical modelling of survey data

First, we fitted independent binomial geostatistical models to baseline prevalence survey data for each of the three STH species (*A. lumbricoides, T. trichiura* and hookworm *spp*.) and used them to predict baseline prevalence at pixel level (5 km resolution). To improve model precision model we explored the use of a set of spatially varying environmental covariates that are known to be potential drivers of STH transmission and included EVI, mean daytime LST, mean night-time LST and soil acidity in the model because they had an approximately linear relationship with prevalence for each of the three species on the log-odds scale (**Supplementary Figures 3-5**). A detailed explanation of the geostatistical modelling process is provided in **Supplementary Section 1** and model parameter estimates are shown in **Supplementary Table 1**, respectively. These true baseline species-specific prevalence surfaces were then used as the input for the two prevalence projecting methods in Step 2.

To create a post-PC benchmark to evaluate the performance of the two projecting methods we followed the same methodology to fit binomial geostatistical models to actual observed post-PC prevalence impact survey data and predict species-specific prevalence surfaces which were then aggregated across species, assuming the risk of an individual contracting each was independent. IU-level prevalence was then classified into five endemicity classes (0-2%, 2-10%, 10-20%, 20-50% and 50-100%), taking into account population density. Model covariate relationship plots and model parameter estimates are included in **Supplementary Figures 6-8** and **Supplementary Table 2**. These predicted outputs are what we try to reproduce using two methods in Step 2.

#### Step 2. Projecting prevalence surface to impact

In Step 2, we projected the baseline prevalence surfaces forward to the time of the proposed impact survey using two different approaches, a multi-state Markov model (12) and a mechanistic transmission model, WORMSIM (13). The mechanistic approach only used the baseline prevalence surface and PC coverage data to achieve this, whereas the Markov model also required midterm prevalence survey data (collected after two rounds of PC in 2015).

#### Method 1: Multi-state Markov model

This method followed a two-stage procedure, described in more detail previously (12). Firstly, we fitted a multi-state Markov model (for each species independently) to baseline and midterm school prevalence data that was grouped into five prevalence categories (0-2%, 2-10%, 10-20%, 20-50% and 50-100%) to estimate regression coefficients for the effect of baseline-midterm PC history on the probability of transition in prevalence category at school-level. We then used these coefficients to predict the transition in endemicity class (also categorised as 0-2%, 2-10%, 10-20%, 20-50% and 50-100%) for each IU between baseline and impact using baseline-impact PC history. Finally, to generate a projected impact prevalence surface we performed a local scaling of the predicted baseline prevalence surface such that the population-weighted mean prevalence of the surface is equal to the endemicity class estimated by the Markov model (on the log-odds scale) and then aggregate across species, assuming independence.

#### Method 2: Mechanistic transmission modelling with WORMSIM

WORMSIM is an established individual-based stochastic model for transmission and control of helminth infections in humans (13), which simulates the life histories of individual humans and individual worms within a closed human population. A formal description of WORMSIM with extensive technical details and mathematical formulae has been published previously (13,14); the main aspects are described in **Supplementary Section 2**. We used WORMSIM to simulate the impact of PC on STH prevalence among SAC in each of the 16 IUs within our study area, based on IU-level baseline prevalence data and PC coverage levels (shown for each IU in **Supplementary Figure 2**). The projected impact prevalence surface was then generated using the same local scaling methodology previously described.

#### Step 3. Simulation study to compare survey design performance

Finally, we conducted a Monte Carlo simulation study to compare the performance of simulated survey designs based on three prevalence surfaces: the two projected impact prevalence surfaces (Step 2) and the benchmark impact surface (Step 1). We evaluated the performance of 24 survey design scenarios in terms of cost and accuracy in determining IU endemicity class (compared to the “true” benchmark prevalence surface at impact from Step 1). The candidate survey designs were created by varying the following: 1) the number of schools to sample - we sampled 20%, 30%, 40%, 60%, 80% and 100% of the total 172 schools used in the original impact survey; 2) the number of children per school - we considered values of 30, 50, 70, and 100.

We then followed the following simulation process. First, for a given design scenario and prevalence surface, the chosen number of schools were randomly sampled from 9,511 georeferenced schools within the study area. Second, prevalence survey data for each STH species was simulated at each school as a realisation of a binomial random variable with probability equal to the predicted prevalence at the school’s location for the given surface and number of trials equal to the number of children per school. Third, three independent binomial geostatistical models were fitted to the simulated school data for each STH species with the same five covariates used in Step 1. Fourth, predicted prevalence surfaces were predicted from the fitted geostatistical models for each of the three species and then combined to create a joint population-weighted ‘any STH’ prevalence surface. Fifth, for each IU, we drew samples from the predictive distribution of the IU-wide population-weighted ‘any STH’ prevalence and calculated the predictive probability of belonging to each of the 5 endemicity classes. The endemicity class with the highest probability was then assigned to the IU. This was repeated 1,000 times for each of the 24 survey designs for each of the three prevalence surfaces.

We then evaluated the performance of each survey design by calculating the proportion of correctly classified IUs. The benchmark for performance for each of the three surfaces was the IU-level endemicity class classified from each projected surface (from which the synthetic school prevalence data was simulated) using a probabilistic classification algorithm for IUs developed previously (6).

## Results

### Multi-state Markov predictions

Prevalence predictions from the multi-state Markov model for each IU and STH species are plotted against the modelled prevalence at impact (our benchmark) in **Supplementary Figure 9**. The Markov model generally performed better for lower prevalence IUs, predicting Hookworm *spp*. and *T. trichiura* prevalence moderately well. For *A. lumbricoides*, however, it significantly underestimated prevalence for the majority of IUs, predicting values in the 0-2% prevalence category for IUs with prevalence rates in excess of 5%.

### WORMSIM predictions

Four different models were developed for each of the species (see **Supplementary Figures 10-15**) and the final models for each species were chosen based on simulated baseline prevalence and expected effectivity of school-based PC in SAC. The selected model for the different species is presented as model #1. For *A. lumbricoides* for IUs with higher baseline prevalence levels, the best fitting model generally predicted a lower impact prevalence than has been observed in the data, suggesting that PC was less effective for decreasing prevalence levels. For *T. trichiura* the predictions of the best fitting model were accurate, closely fitting values observed in the data at impact in 12 out of 16 IUs. For hookwork *spp*., the best fitting model was able to predict impact prevalence of hookworm *spp*. relatively accurately. However, the benchmark impact prevalence levels were always in the lower range of the predicted fluctuations in prevalence over time, the opposite of what would be expected (that impact prevalence would be on the higher ends of the ranges) due to sampling taking place directly before the next round of PC took place.

### Projected prevalence surfaces at impact

Projected prevalence surfaces for the Markov and mechanistic models were generated by scaling the baseline prevalence surface by each model’s IU-level prevalence predictions. The projected surfaces of *A. lumbricoides* prevalence at impact for both the Markov and mechanistic models **(Supplementary Figure 16**) failed to capture the hotspots above 10% prevalence in Nyanza and Rift Valley regions (see **Figure 1A** for study area map), although the mechanistic projected surface performed better in this area, predicting prevalence in the range of 5-10%. In contrast, the Markov projected surface consistently predicted prevalence in the range of 0-1% for this area. This is likely to be because prevalence in these areas did not decline significantly between baseline and impact, suggesting a limited impact of PC. The projected prevalence surfaces for both models capture the low prevalence areas (0-5%) well, but the mechanistic projected surface slightly overestimated prevalence in the very low (0-1%) prevalence Coast region.

For *T. trichiura*, the Markov projected surface generally underestimated prevalence and missed the hotspots in the Nyanza and Rift Valley regions (**Supplementary Figure 17)**. In contrast, the mechanistic projected surface captured hotspots but tended to overestimate prevalence in the lowest prevalence areas of the Rift Valley. Projected surfaces for both models overestimated prevalence in the Coast region where the predictions from the geostatistical model at impact indicated that prevalence had fallen to very low levels (0-1%).

For hookworm *spp*., the projected surfaces for both models performed well, although the mechanistic projections overestimated prevalence in the Coast, Nyanza and Western regions (**Supplementary Figure 18**). The improved performance for the projected surfaces of both models for hookworm *spp*. relative to the other two species appeared to be driven by the consistent reduction in prevalence between baseline and impact across all IUs with an absence of any persistent hotspots.

Both the Markov and mechanistic projected surfaces captured the reduction in STH prevalence in the three IUs in the Coast region, although they both slightly overestimated prevalence in the western IU in the region. The high prevalence area (20-60%) in the middle of the Nyanza and Rift valley regions was missed by both models, although the mechanistic projected surface predicted slightly higher prevalence (10-30%) than the Markov surface (0-20%). The Markov projected surface underestimated the prevalence in the Western region (2-5%), whereas the mechanistic projections more accurately predicted the true prevalence in this area.

The Markov projected surface predicted *A. lumbricoides* and *T. trichiura* prevalence in the Coast and Nyanza regions accurately (**Figure 2A**), but consistently predicted lower prevalence values in the Rift Valley region and some areas of the Western region. For hookworm *spp*. the regional differences were less profound with some areas of the Coast and Nyanza regions having lower predicted values than the benchmark values. The mechanistic projected surface predicted *A. lumbricoides* and *T. trichiura* prevalence at impact more accurately at lower prevalence levels (**Figure 2B**), but generally tended to underestimate prevalence at higher modelled prevalence levels. However, in the Coast region the mechanistic projections overestimated *T. trichuria* prevalence. For hookworm *spp*., the mechanistic projections consistently overestimated prevalence, with this most pronounced in the Western region.

### Survey simulation results

There was significant variation in the outcome across simulations for each survey design. **Figure 3** shows how the proportion of IUs that were correctly classified in terms of endemicity class increased with a higher number of children sampled in each school and a higher proportion of schools sampled. Compared to the performance of simulated surveys based on the best available information (i.e., the predicted surface from the geostatistical model fitted to the impact data), surveys based on the mechanistic and Markov model projected surfaces generally performed worse. The performance of surveys based on the Markov projected surface was consistently between 10-20 percentage-points lower, while the performance of survey designs based on the mechanistic model were similar to or at most 7 percentage-points lower than the benchmark.

**Figure 3.**
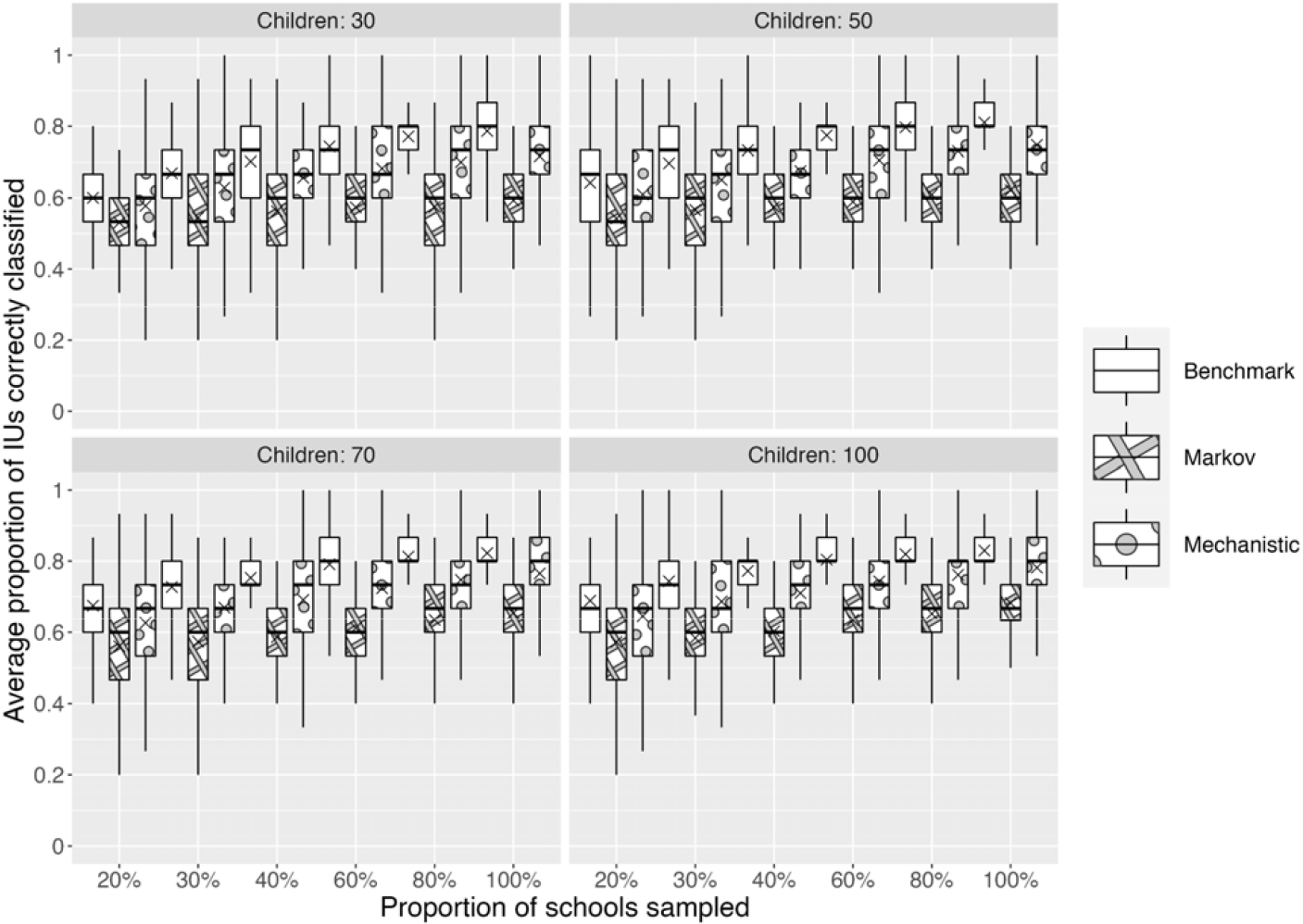
Comparison of the performance (percentage of IUs with the endemicity level correctly classified) of a range of survey designs (for varying values of the number of children per school and proportion of schools sampled) for the benchmark geostatistical model predictions from the impact survey data (i.e., the best available information), and the Markov and mechanistic projected surfaces.

## Discussion

In this study we compared two approaches for projecting STH prevalence at impact that integrate model-based geostatistical predictions of baseline prevalence and model-based forward predictions of prevalence using i) WORMSIM, a mechanistic transmission model, and ii) multi-state Markov models for school-level prevalence categories. We then evaluated their performance using STH prevalence data from Kenya. This is the first study to have directly compared a mechanistic and a more empirical Markov model for projecting prevalence during PC in the context of guiding decisions on IAS design.

We found that while the Markov and mechanistic approaches both generally performed well for projecting STH prevalence at impact, prediction accuracy was lower in areas with persistent high prevalence hotspots that did not reduce significantly after PC; these were mostly concentrated in the Rift Valley. While the mechanistic approach was less prone to this than the Markov approach, both models underestimated prevalence in these areas because of the limited impact of PC on prevalence compared with the rest of the study area where PC was observed to significantly reduce prevalence. This observed variation in PC effectiveness within the study area may be a result of measurement error in the PC coverage data or in the baseline and impact prevalence survey data. Interestingly, this was not the case for hookworm *spp*., with prevalence consistently declining following PC in all IUs, and consequently both models predicted prevalence at impact accurately. The local scaling approach used to project proxy prevalence surfaces at impact by scaling baseline prevalence predictions from the geostatistical model by IU-level projections assumed that the spatial distribution of predictions at impact was conditional on the spatial distribution at baseline. We found that for our case study this was a good approximation, but in future applications it will perform less well in areas where there are abrupt changes in the spatial variation in prevalence within an IU between baseline and impact, e.g., due to high spatial variation in PC uptake.

In the context of survey design, and in statistical design more generally, any sample size calculation must be made on the basis of assumptions that represent a best guess at the true state of the natural process under investigation. The better the guess, the more likely the chosen design will deliver the required precision whilst avoiding wasteful over-sampling, but this can never be guaranteed. Our projected prevalence surfaces at impact were reasonably well-calibrated against geostatistical predictions using the impact survey data, albeit with considerable uncertainty and with some exceptions, most notably with respect to some species in the Rift Valley region where the Markov approach performed particularly badly. Despite these challenges, our simulation study demonstrated that the projected surfaces from the mechanistic approach and, to a lesser degree, from the Markov approach, were highly informative for guiding survey design as measured by the proportion of correctly classified IUs. Given the budget constraints that apply in regions where STH is prevalent, both the design of an IAS and the subsequent analysis of IAS data should be conducted as efficiently as possible. Our simulation study only considered spatially random sampling, but further improvements in the efficiency of a survey design can sometimes be achieved by spatially regulated sampling, as has been shown previously (6).

For the design of future impact surveys in other geographical areas, the Markov model is limited by its reliance on midterm data to estimate the effect of PC. In the absence of midterm survey data, the Markov model could still be applied using parameters for PC efficacy estimated from this Kenya case study. However, in this case the mechanistic model, which does not require midterm data, is likely to perform better and therefore to be more generalisable to other geographical areas because it explicitly models the interaction between PC and STH transmission dynamics.

This study demonstrated that the mechanistic approach more accurately projected prevalence at impact and provided more accurate information for guiding survey design. The usefulness of the two approaches for projecting and survey design considered in this study is not confined to STH. The Markov model is not disease-specific and can be applied directly to other NTDs that are controlled with PC. The mechanistic model used here is an STH-specific transmission model and would need to be replaced with a validated transmission model for any other NTD of interest. Our results suggest that if a validated transmission model is available, it should be used to guide survey design. For both approaches, environmental explanatory variables that are known to be predictors of prevalence for the NTD of interest should be included in the geostatistical models used for predicting prevalence at baseline.

## Data Availability

All human data used in this analysis are publicly available via the Global Atlas of Helminth Infections (https://www.thiswormyworld.org/) and the ESPEN portal (https://espen.afro.who.int/). All environmental data used are publicly available from MODIS and WorldPop.

## Supplementary material

### Supplementary Section 1: Geostatistical modelling framework

#### S1.1 The geostatistical model

We denote by *P*(*x*) the prevalence of STH at location *x*. Our model for the variation in *P*(*x*) throughout the region of interest is that

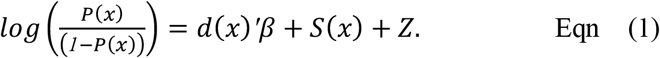

In Equation (1), *d*(*x*) is a vector of covariates associated with regression coefficients β. This component of the model accounts for variation in prevalence that can be explained by measured characteristics of the location *x*; see Table S3. The terms *S*(*x*) and *Z* account for any remaining variation that cannot be explained by measured characteristics of *x*. The term *S*(*x*)is a spatially correlated Gaussian process with mean zero and covariance structure

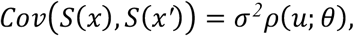

where *u =* ‖*x* − *x′*‖ is the Euclidean distance between *x* and *x*^*′*^, σ^*2*^ is the variance and

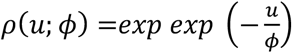

is the correlation between *S*(*x*) and *S*(*x′*). The term *Z* in Equation (1) is a Normally distributed random variable with zero mean and variance τ^*2*^ that varies independently between locations; it accounts for variation in unmeasured characteristics of the sampled individuals that affect their personal exposure to STH.

We denote by *x*_*1*_, …, *x*_*n*_ the set of sampled locations. Conditional on *P*(*x*_*i*_), the numbers *Y*_*i*_ of individuals who test positive out of *m*_*i*_ sampled individuals at *x*_*i*_ are independent binomially distributed random variables, with binomial probabilities *P*(*x*_*i*_) and denominators *m*_*i*_.

#### S1.2 Parameter estimation

We carry out parameter estimation using Monte Carlo Maximum Likelihood (MCML), implemented in PrevMap, an R package for analysing prevalence data, freely available from the Comprehensive R Archive Network (*www*.*r-project*.*org*).

Let 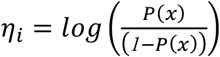. The joint conditional density of *Y=Y_1_*,…,*Y_n_* is 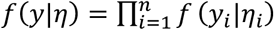.

The likelihood function for the set of model parameters *ψ* is obtained by integrating out the random components*S*(*x*_*i*_)and*Z*_*i*_ from η_*i*_, hence

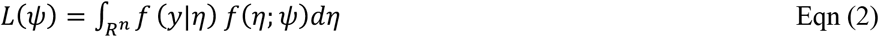

Where *f* (η; *ψ*)is a multivariate Normal density.

To approximate the integral in Equation 2 we use a Markov Chain Monte Carlo (MCMC) algorithm to generate a sample η_(*1*)_,..., η_(*N*)_ from the conditional distribution of η given *y* and approximate the likelihood as

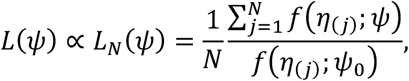

Where *ψ*_*0*_ is our best guess for the initial parameter values.

#### S1.3 Prediction

Here, we use plug-in prediction, meaning that we use the Monte Carlo maximum likelihood parameter estimate *ψ*^ in place of the unknown *ψ*.

Our goal is to predict prevalence throughout the region of interest, *A*. We approximate this by a regular grid of points *x*_*n*+*1*_, …, *x*_*n*+*q*_ that cover *A*. Our predictive target is the set of values

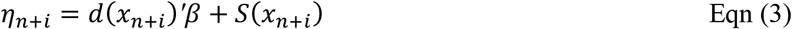

Note that Equation 3 excludes the term *Z* in Equation 1, which relates to characteristics of the sampled individuals at a location rather than of the location itself.

The *predictive distribution* of η^*^ *=* (η_*n*+*1*_, …, η_*n*+*q*_) is its conditional distribution given *y*,

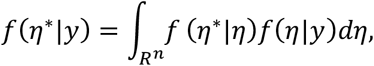

where we have used the fact that η^*^ and *y* are conditionally independent given η. It follows that to generate a sample from the predictive distribution of η^*^ we first sample from *f*(η|*y*) and then from*f*(η^*^|η). A sample from the joint predictive distribution of prevalence throughout *A* follows by direct transformation, using the formula

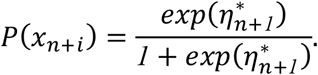

Our point prediction of prevalence at any location *x* is the mean of the sampled values of *P*(*x*). The predictive probability that prevalence lies within any stated range, say *c* to *d*, is the proportion of sampled values that lie between *c* and *d*. The predictive distribution of prevalence at the implementation unit level, *P*_*IU*_, is computed as a population-weighted average of the pixel level prevalence *P*(*x*), hence

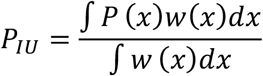

where *w*(*x*) is an estimate of the population at location *x* obtained from WorldPop (*https://www.worldpop.org/*) and the integral is over the whole of the IU.

### Supplementary Section 2: Description of WORMSIM

WORMSIM simulates the life histories of individual humans and individual worms within a closed human population. The population-level age- and sex-distribution are based on pre-specified fertility rates and life tables. A comparison of the population distribution for WORMSIM and the Kenyan population is provided in (see **Supplementary Figure 2.1** below).

**Supplementary Figure 2.1.**
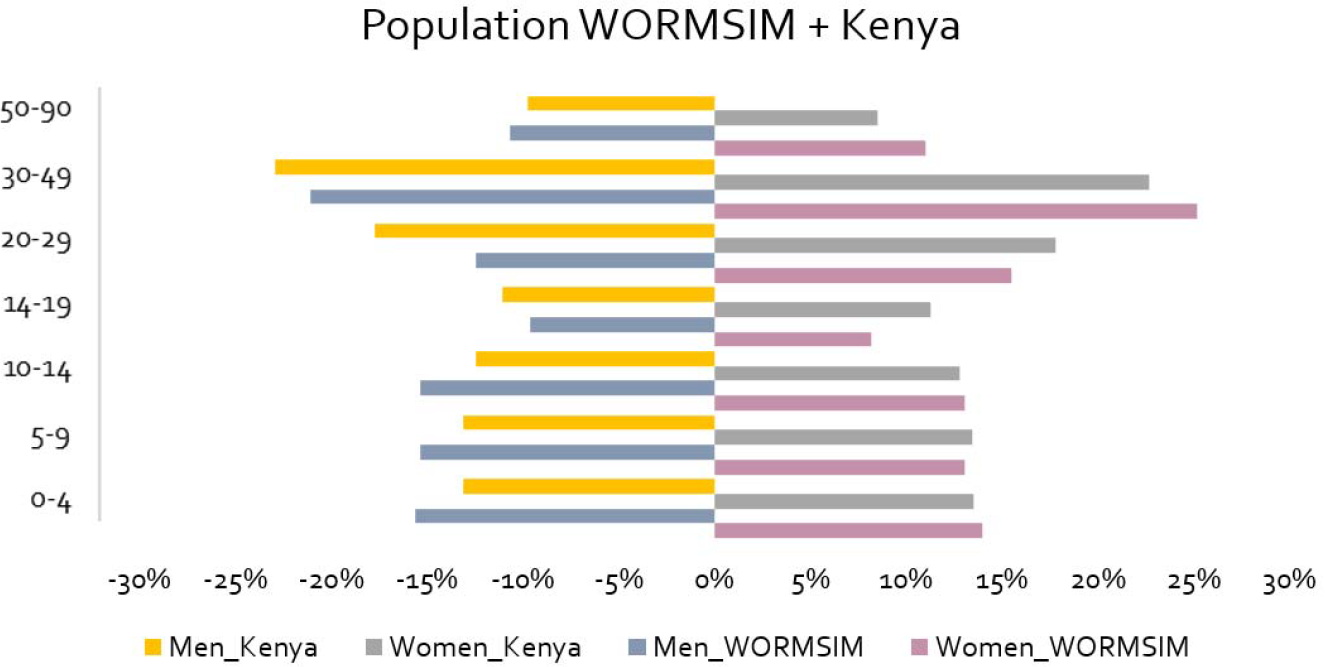
Population age pyramid by sex for WORMSIM population and Kenyan population.

The prevalence and intensity distribution of worm infections is determined by model parameters for the transmission rate (overall rate and differences by age) and level of exposure heterogeneity within a population. For this study, we assume standard level of exposure heterogeneity for each species (gamma distribution with shape parameter *k* (13)), and we adapted the overall transmission rate for each IU to replicate IU-level baseline prevalences of infection as estimated from the NSBDP data. The overall transmission rate was calibrated following two different assumptions: transmission increasing with age versus transmission being independent of age (13,15). However, in another widely applied individual-based stochastic transmission model by Imperial College London (ICL) (15), the modellers assume that age-dependent contribution to infection is proportional to age-dependent exposure.

For *T. trichiura* and *A. lumbricoides*, we explored two different values for (species-specific) exposure heterogeneity (= p1), to reflect different potential exposure and transmission dynamics. Simulating stable transmission dynamics with very low overall prevalence levels is known to be challenging with WORMSIM (13,16). Therefore, if baseline prevalence levels were lower than a certain value, we ran the model simulations for a higher cut-off prevalence value (for hookworm *spp*. 20%; for *T. trichiura* 30%; and for *A. lumbricoides* 25%) and scaled the predicted prevalence values back to the measured lower baseline prevalence.

The IU-level baseline prevalences to which WORMSIM was calibrated were estimated for each IU by weighting the baseline geostatistical model prevalence prediction surface by population density and calculating two IU-level summary statistics: mean prevalence and prevalence tertiles. Prevalence tertiles were created by taking the grid-level population weighted-prevalence divided into three ordered groups. WORMSIM further simulates the impact of PC on STH infection levels, assuming that treatment kills a proportion of adult worms (efficacy varying by species and drug as in previous studies (13)) and accounting for the proportion of the population that takes up PC. For this study, we adopt coverage levels directly from the data, assuming mixed compliance. For simulating infection with hookworm *spp*., we explored two different functions for infection exposure by age, further illustrated in a previous comparison study (17). In WORMSIM we originally assume that the contribution and exposure to transmission increases with age up to the age of ten, after which the practice of defecation is assumed to remain similar regardless of age. A similar pattern is applied to *T. trichiura* and *A. lumbricoides*.

For each species, we simulated different model scenarios for different levels of exposure heterogeneity, age-dependent exposure patterns and baseline (**Supplementary Table 2.1** and **Supplementary Figures 9-11**), and selected the most appropriate scenario based on the expected effectivity of school-based PC in SAC as reported in the scientific literature (16). For all three species, we simulated prevalence levels over time based on each of the aforementioned prevalence summary statistics i) IU-level mean population-weighted prevalence and ii) IU-level population-weighted prevalence tertiles to account for real-life spatial heterogeneity in prevalence that might influence infection dynamics.

To generate a proxy impact surface, local scaling of the predicted baseline prevalence surface was then conducted using the prevalence simulations at impact generated by WORMSIM for each IU following the methodology described in ‘Local scaling to generate a proxy impact surface’.

The predicted *A. lumbricoides* prevalence using four different model scenarios is provided in **Supplementary Figure 10**. The predicted prevalence from the final model (model #1) and measured baseline and impact prevalence levels are displayed in **Supplementary Figure 11**. The predicted *T. trichiura* prevalence using 4 different model scenarios is provided in **Supplementary Figure 12**. The predicted prevalence from the final model (model #1) and measured baseline and impact prevalence levels are displayed in **Supplementary Figure 13**. The predicted hookworm spp. prevalence using different model scenarios is provided in **Supplementary Figure 14**. The predicted prevalence from the final model (model #1) and measured baseline and impact prevalence levels are displayed in **Supplementary Figure 15**.

## Supplementary Figures & Tables

**Supplementary Table 2.1.** Overview of explored WORMSIM models shown for each STH species.

|  |  | *A. lumbricoides* | *T. trichiura* | Hookworm spp. |
| --- | --- | --- | --- | --- |
| **Model 1** | Exposure heterogeneity | High exposure (p1 = 0.8) | High exposure (p1 = 0.35) | High exposure (p1 = 0.35) |
|  | Age-exposure function | Assumption of transmission increasing with age (EMC model) | Assumption of transmission increasing with age (EMC model) | Assumption of transmission being independent of age (ICL model) |
|  | Baseline prevalence | Based on tertiles | Based on tertiles | Based on tertiles |
| **Model 2** | Exposure heterogeneity | High exposure (p1 = 0.8) | High exposure (p1 = 0.35) | High exposure (p1 = 0.35) |
|  | Age-exposure function | Assumption of transmission increasing with age (EMC model) | Assumption of transmission increasing with age (EMC model) | Assumption of transmission being independent of age (ICL model) |
|  | Baseline prevalence | Overall mean | Overall mean | Overall mean |
| **Model 3** | Exposure heterogeneity | Low exposure (p1 = 0.4) | Low exposure (p1 = 0.2) | High exposure (p1 = 0.35) |
|  | Age-exposure function | Assumption of transmission increasing with age (EMC model) | Assumption of transmission increasing with age (EMC model) | Assumption of transmission increasing with age (EMC model) |
|  | Baseline prevalence | Based on tertiles | Based on tertiles | Based on tertiles |
| **Model 4** | Exposure heterogeneity | Low exposure (p1 = 0.4) | Low exposure (p1 = 0.2) | High exposure (p1 = 0.35) |
|  | Age-exposure function | Assumption of transmission increasing with age (EMC model) | Assumption of transmission increasing with age (EMC model) | Assumption of transmission increasing with age (EMC model) |
|  | Baseline prevalence | Overall mean | Overall mean | Overall mean |

**Supplementary Figure 1.**
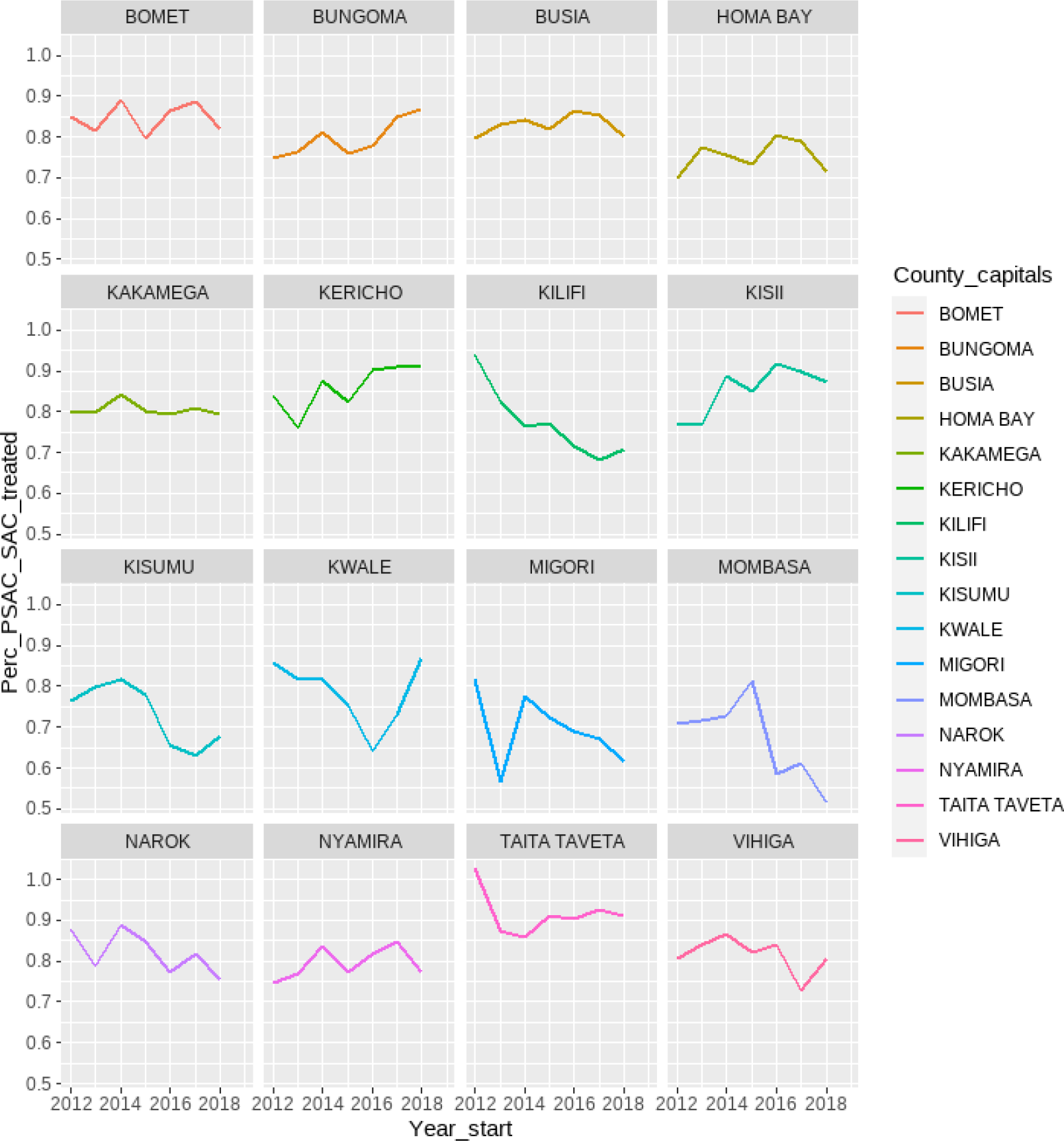
Preventive chemotherapy (PC) coverage data between baseline and impact surveys for the 16 IUs in Southwest Kenya. Data was obtained through Evidence Action.

**Supplementary Figure 2.**
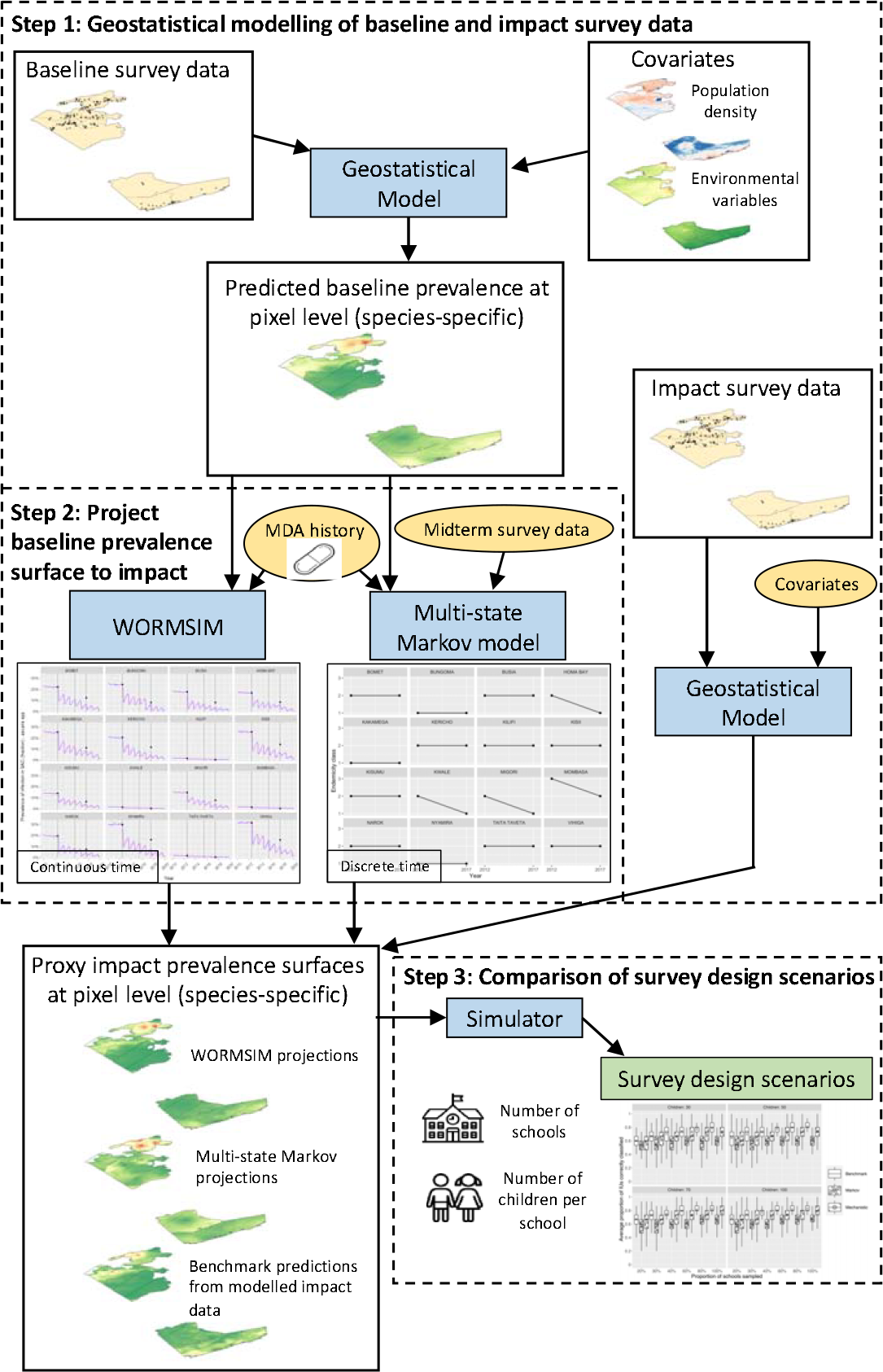
Overview of the study modelling process

**Supplementary Figure 3.**
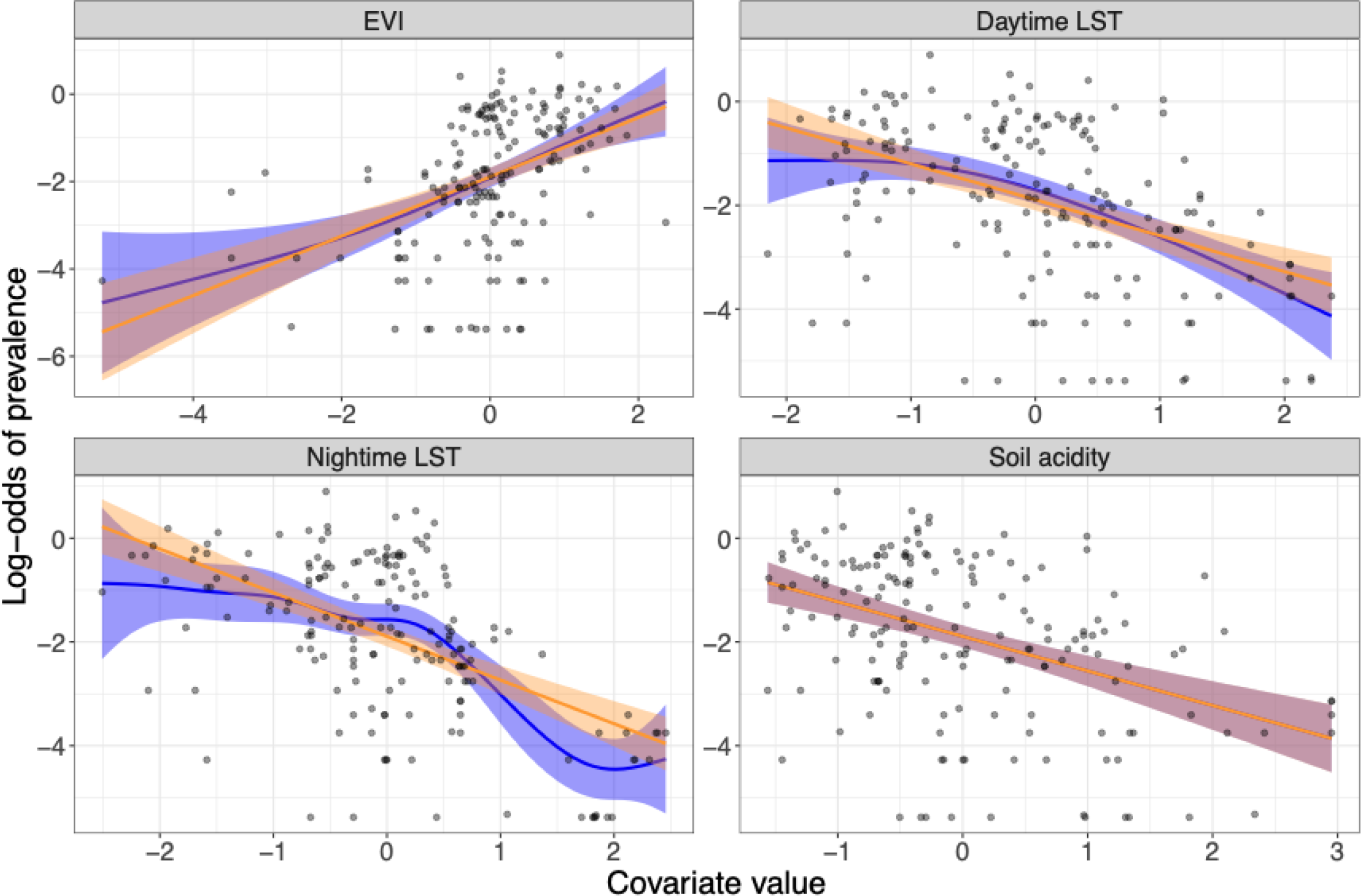
Generalized Additive Model (GAM) and linear model dependence plots for the log-odds of *A. lumbricoides* prevalence at baseline plotted against the continuous environmental covariates considered in this analysis (data are plotted as points and shaded areas correspond to 95% confidence intervals): EVI (Enhanced Vegetation Index), mean daytime LST (land surface temperature), mean nighttime LST, soil acidity.

**Supplementary Figure 4.**
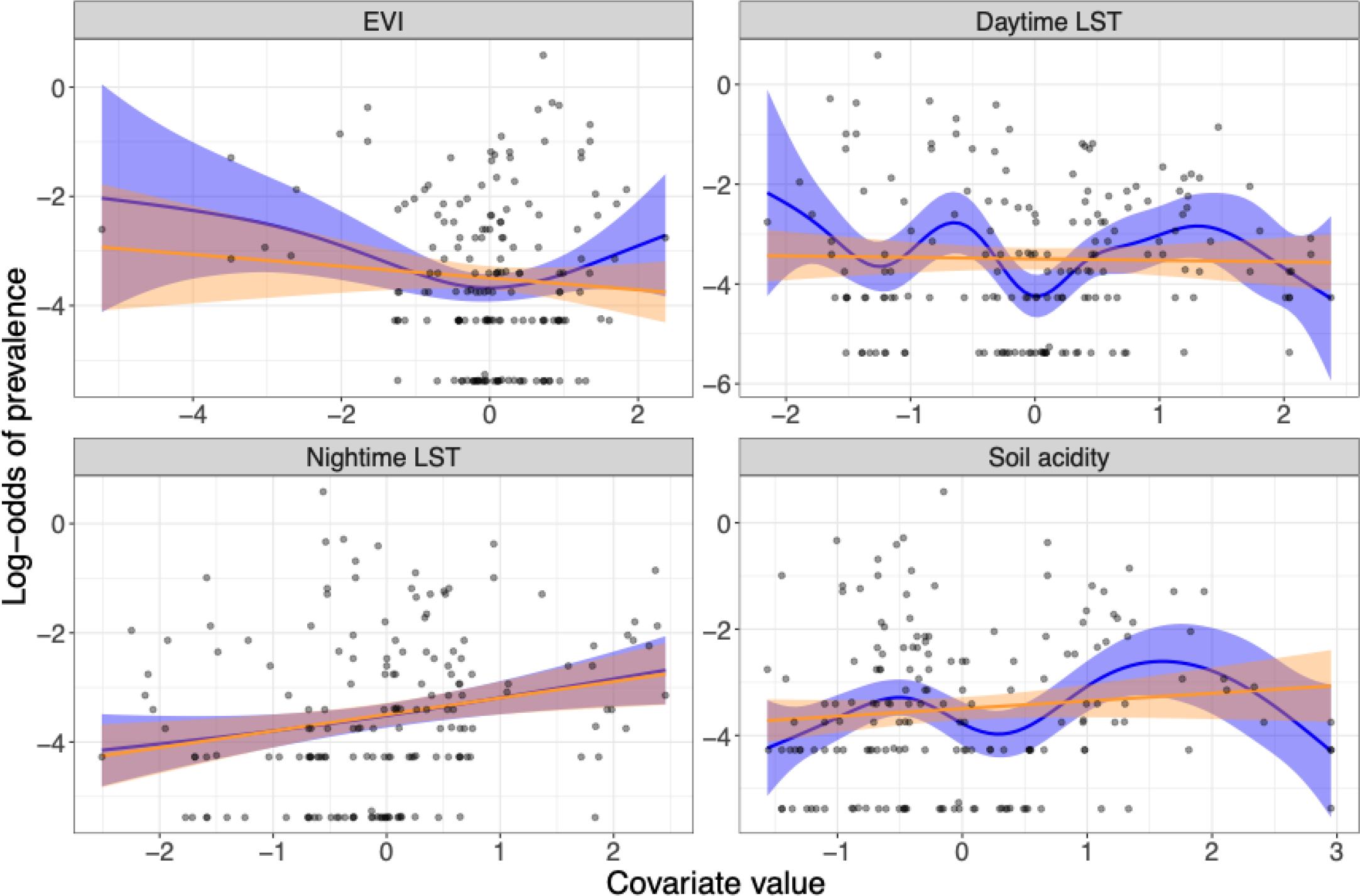
Generalized Additive Model (GAM) and linear model dependence plots for the log-odds of *T. trichiura* prevalence at baseline plotted against the continuous environmental covariates considered in this analysis (shaded areas correspond to 95% confidence intervals): EVI (Enhanced Vegetation Index), mean daytime LST (land surface temperature), mean nighttime LST, soil acidity.

**Supplementary Figure 5.**
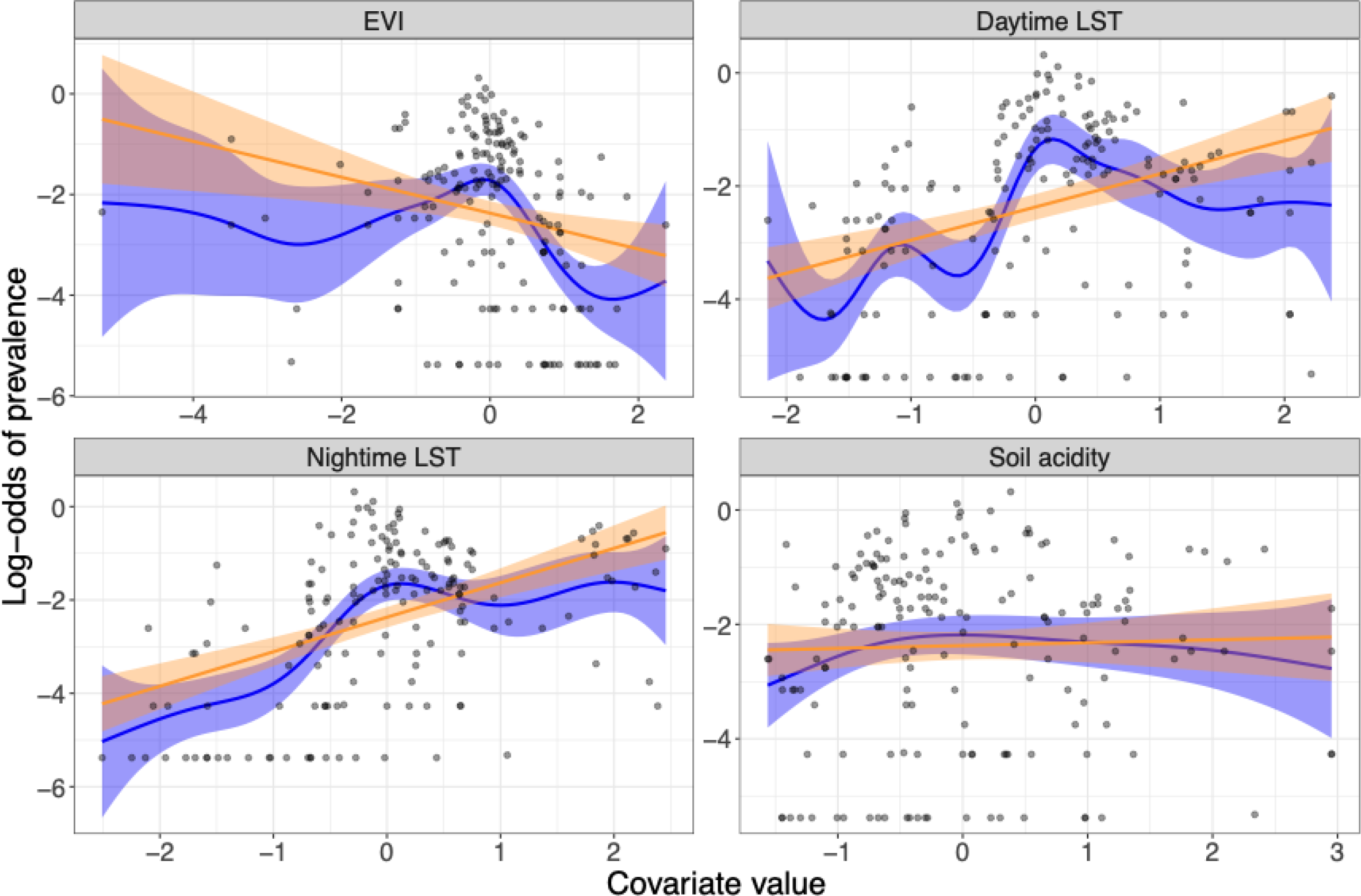
Generalized Additive Model (GAM) and linear model dependence plots for the log-odds of hookworm *spp*. prevalence at baseline plotted against the continuous environmental covariates considered in this analysis (shaded areas correspond to 95% confidence intervals): EVI (Enhanced Vegetation Index), mean daytime LST (land surface temperature), mean nighttime LST, soil acidity.

**Supplementary Figure 6.**
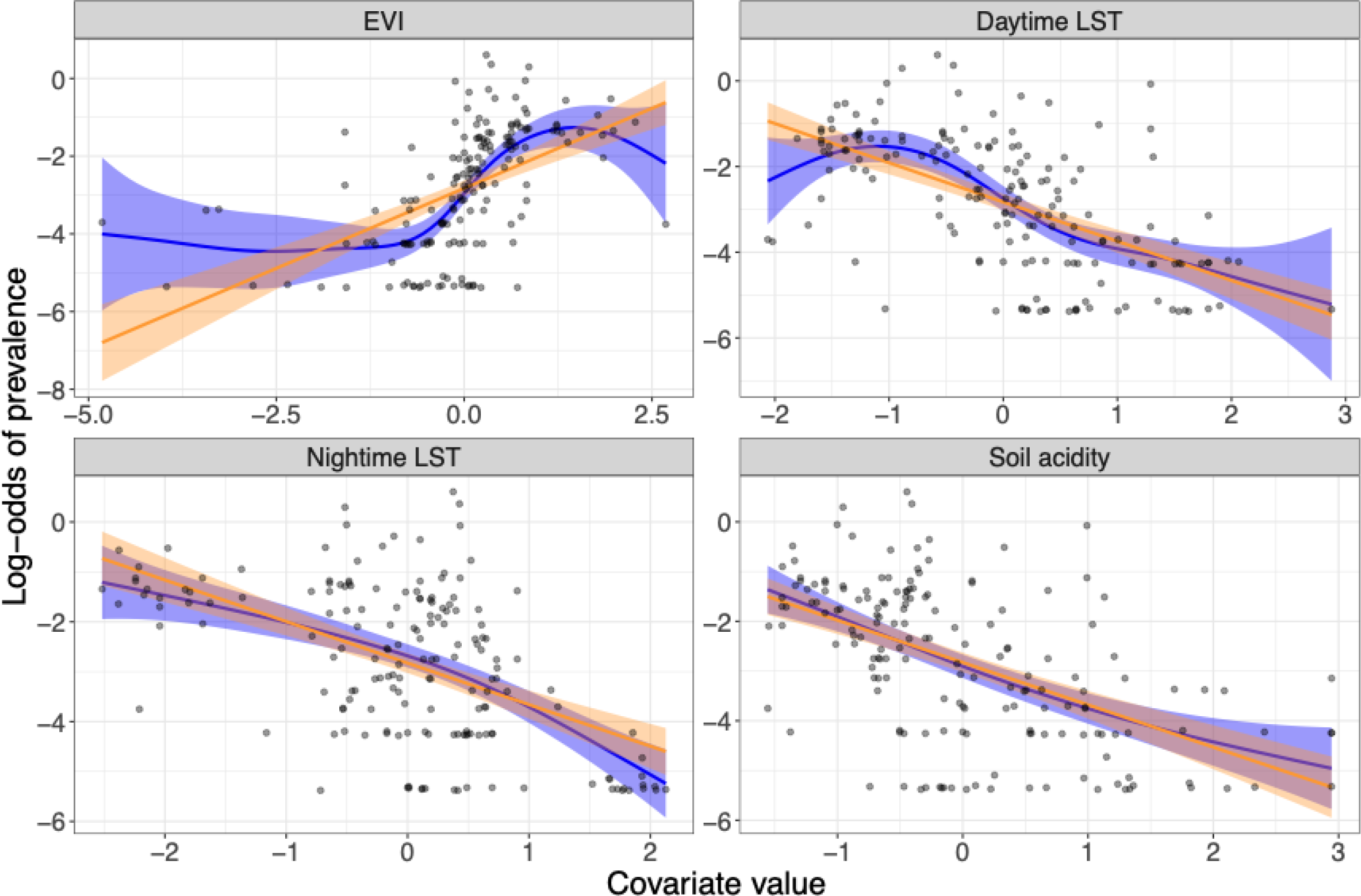
Generalized Additive Model (GAM) and linear model dependence plots for the log-odds of *A. lumbricoides* prevalence at impact plotted against the continuous environmental covariates considered in this analysis (data are plotted as points and shaded areas correspond to 95% confidence intervals): EVI (Enhanced Vegetation Index), mean daytime LST (land surface temperature), mean nighttime LST, soil acidity.

**Supplementary Table 1.** Monte Carlo maximum likelihood estimates and corresponding 95% confidence intervals for the baseline geostatistical model (continuous covariates were standardised).

|  | *A. lumbricoides* | *T. trichiura* | Hookworm *spp.* |
| --- | --- | --- | --- |
| EVI | 0.265 (-0.105, 0.635) | 0.018 (-0.392, 0.428) | 0.279 (-0.070, 0.628) |
| LST (day) | -0.101 (-0.502, 0.299) | -0.161 (-0.683, 0.362) | 0.360 (-0.070, 0.790) |
| LST (night) | -0.291 (-0.794, 0.213) | 0.252 (-0.658, 1.162) | 0.748 (0.126, 1.371) |
| Soil acidity | 0.013 (-0.401, 0.426) | -0.049 (-0.589, 0.491) | -0.286 (-0.717, 0.145) |
| Region - Coast (ref) | - | - | - |
| Region - Nyanza | 2.346 (1.189, 3.502) | -1.191 (-4.107, 1.726) | 1.126 (-0.403, 2.656) |
| Region - Rift valley | 2.137 (0.800, 3.475) | 1.446 (-1.312, 4.203) | -0.435 (-2.090, 1.220) |
| Region - Western | 2.842 (1.755, 3.929) | -0.561 (-3.165, 2.043) | 1.462 (-0.042, 2.965) |
| $\sigma^2$ | 0.726 (0.395, 1.333) | 1.967 (0.742, 5.210) | 0.865 (0.374, 1.998) |
| $\phi$ (km) | 8.517 (3.598, 20.161) | 53.296 (14.070, 201.885) | 24.356 (6.167, 96.197) |
| $\tau^2$ | 0.372 (0.130, 2.024) | 0.749 (0.120, 1.206) | 0.550 (0.209, 1.936) |

**Supplementary Figure 7.**
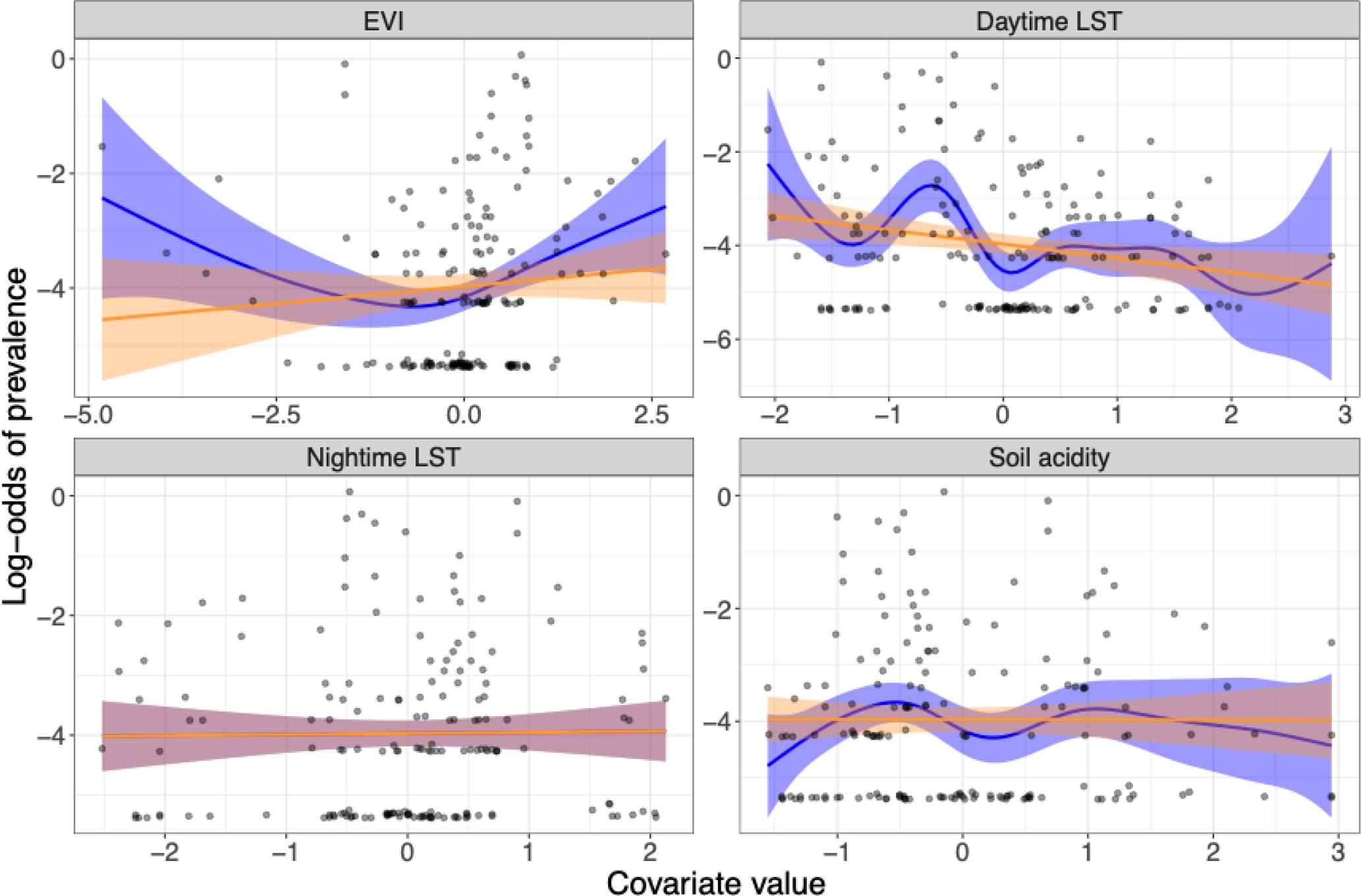
Generalized Additive Model (GAM) and linear model dependence plots for the log-odds of *T. trichiura* prevalence at impact plotted against the continuous environmental covariates considered in this analysis (data are plotted as points and shaded areas correspond to 95% confidence intervals): EVI (Enhanced Vegetation Index), mean daytime LST (land surface temperature), mean nighttime LST, soil acidity.

**Supplementary Figure 8.**
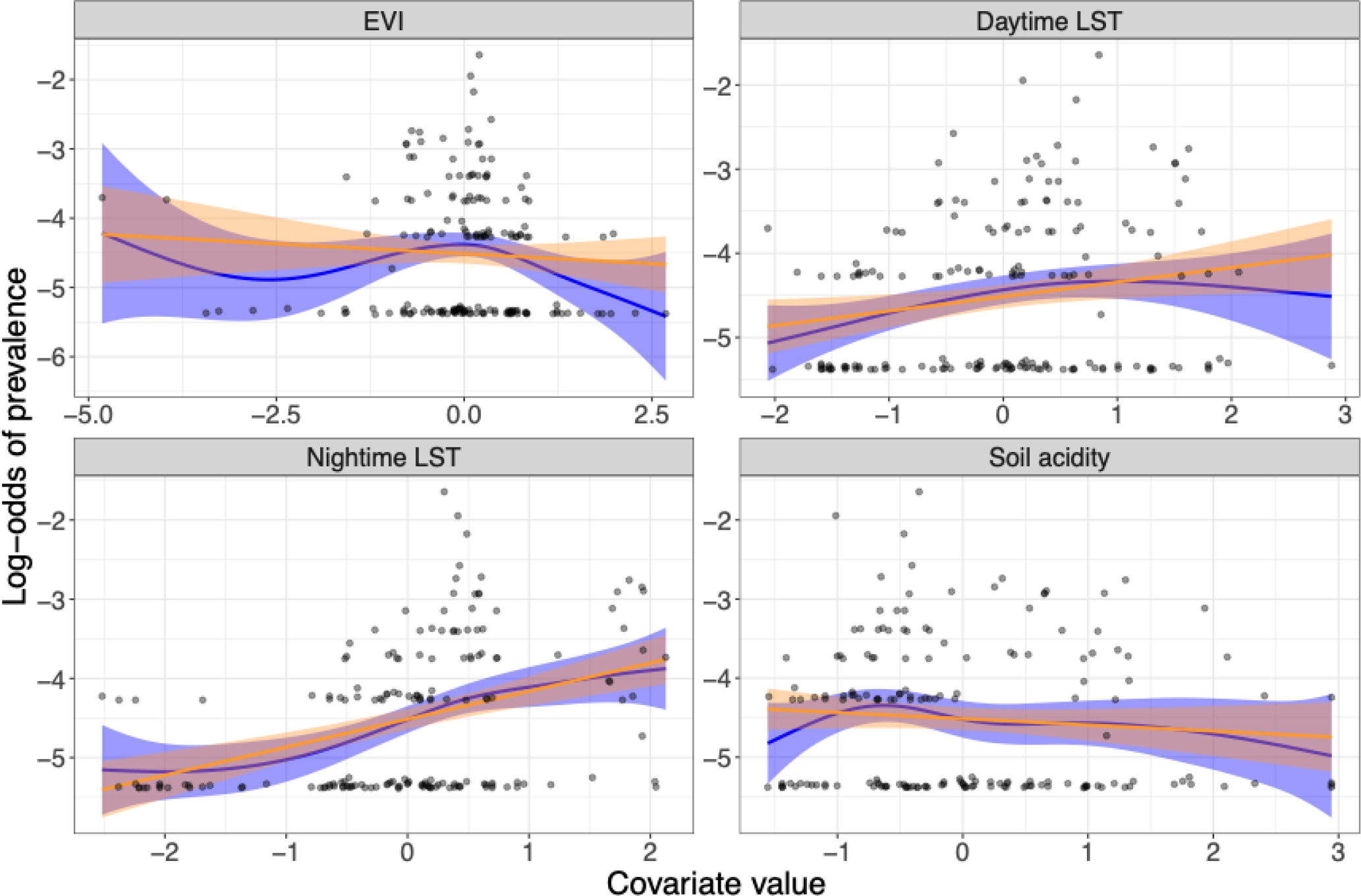
Generalized Additive Model (GAM) and linear model dependence plots for the log-odds of hookworm *spp*. prevalence at impact plotted against the continuous environmental covariates considered in this analysis (data are plotted as points and shaded areas correspond to 95% confidence intervals): EVI (Enhanced Vegetation Index), mean daytime LST (land surface temperature), mean nighttime LST, soil acidity.

**Supplementary Table 2.** Monte Carlo maximum likelihood estimates and corresponding 95% confidence intervals for the baseline geostatistical model (continuous covariates were standardised).

|  | *A. lumbricoides* | *T. trichiura* | Hookworm *spp.* |
| --- | --- | --- | --- |
| EVI | 0.565 (0.158, 0.972) | 0.347 (-0.069, 0.763) | 0.290 (0.046, 0.533) |
| LST (day) | -0.642 (-1.069, -0.215) | -0.746 (-1.321, -0.172) | 0.191 (-0.080, 0.463) |
| LST (night) | 0.386 (-0.132, 0.904) | 1.276 (0.363, 2.188) | 1.328 (0.962, 1.693) |
| Soil acidity | -0.079 (-0.522, 0.364) | 0.090 (-0.483, 0.662) | -0.575 (-0.870, -0.279) |
| Region - Coast (ref) | - | - | - |
| Region - Nyanza | 5.003 (2.941, 7.064) | 1.005 (-1.941, 3.951) | 1.158 (0.337, 1.979) |
| Region - Rift valley | 5.057 (2.971, 7.143) | 4.149 (1.384, 6.914) | 0.750 (-0.240, 1.740) |
| Region - Western | 5.019 (3.029, 7.009) | 0.933 (-1.481, 3.346) | 0.988 (0.268, 1.708) |
| $\sigma^2$ | 0.903 (0.566, 1.439) | 1.708 (0.574, 5.086) | 0.559 (0.389, 0.803) |
| $\phi$ (km) | 6.530 (3.271, 13.040) | 38.231 (8.182, 178.633) | 8.518 (5.063, 14.329) |
| $\tau^2$ | 0.145 (0.022, 1.171) | 0.602 (0.089, 1.395) | 0.041 (0.013, 0.422) |

**Supplementary Figure 9.**
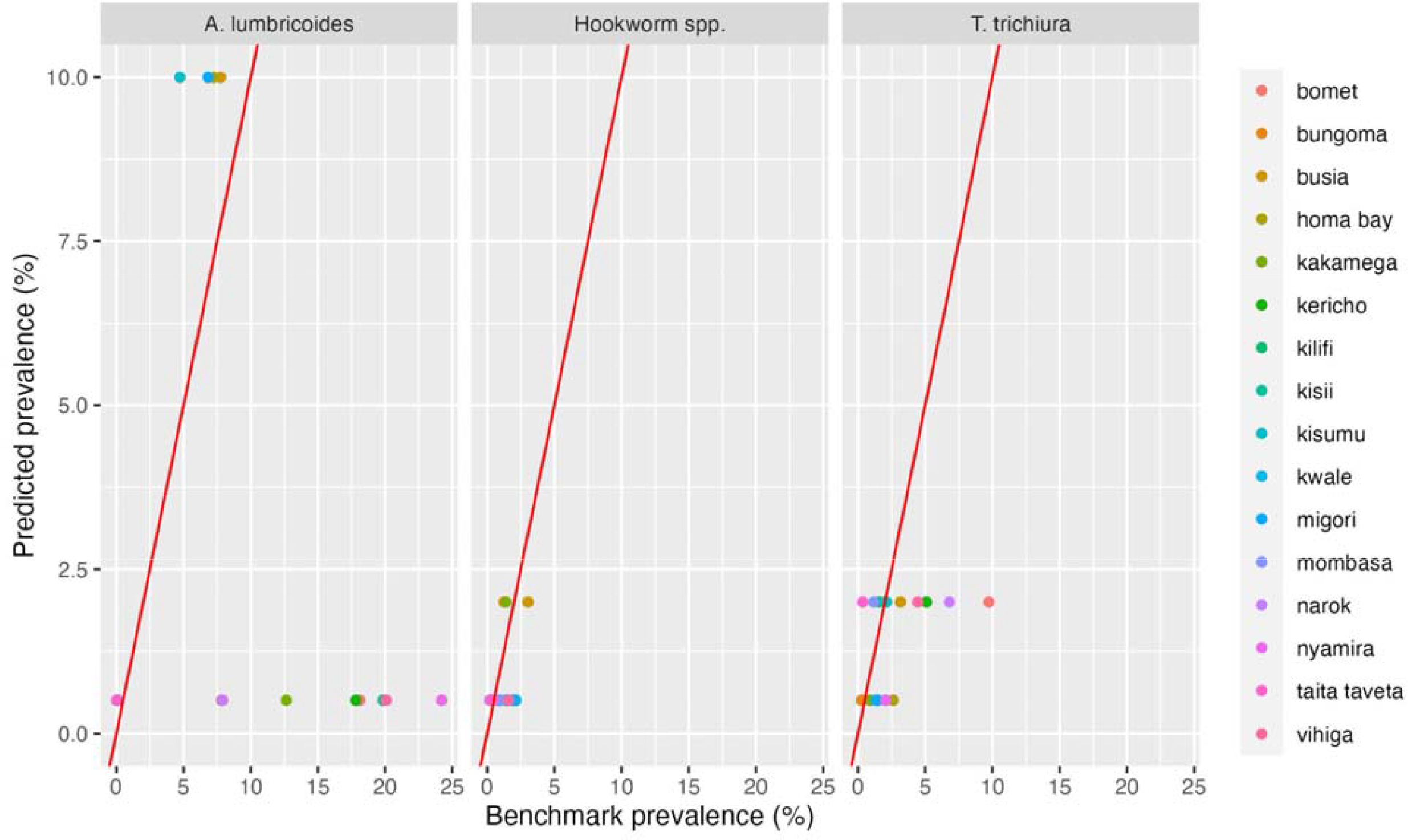
Predicted prevalence for the Markov model for i) *A. lumbricoides*, ii) *Hookworm spp*. And iii) *T. trichiura* among school-aged children (SAC) at 16 IUs in Southwest Kenya, plotted against the benchmark prevalence from the impact survey.

**Supplementary Figure 10.**
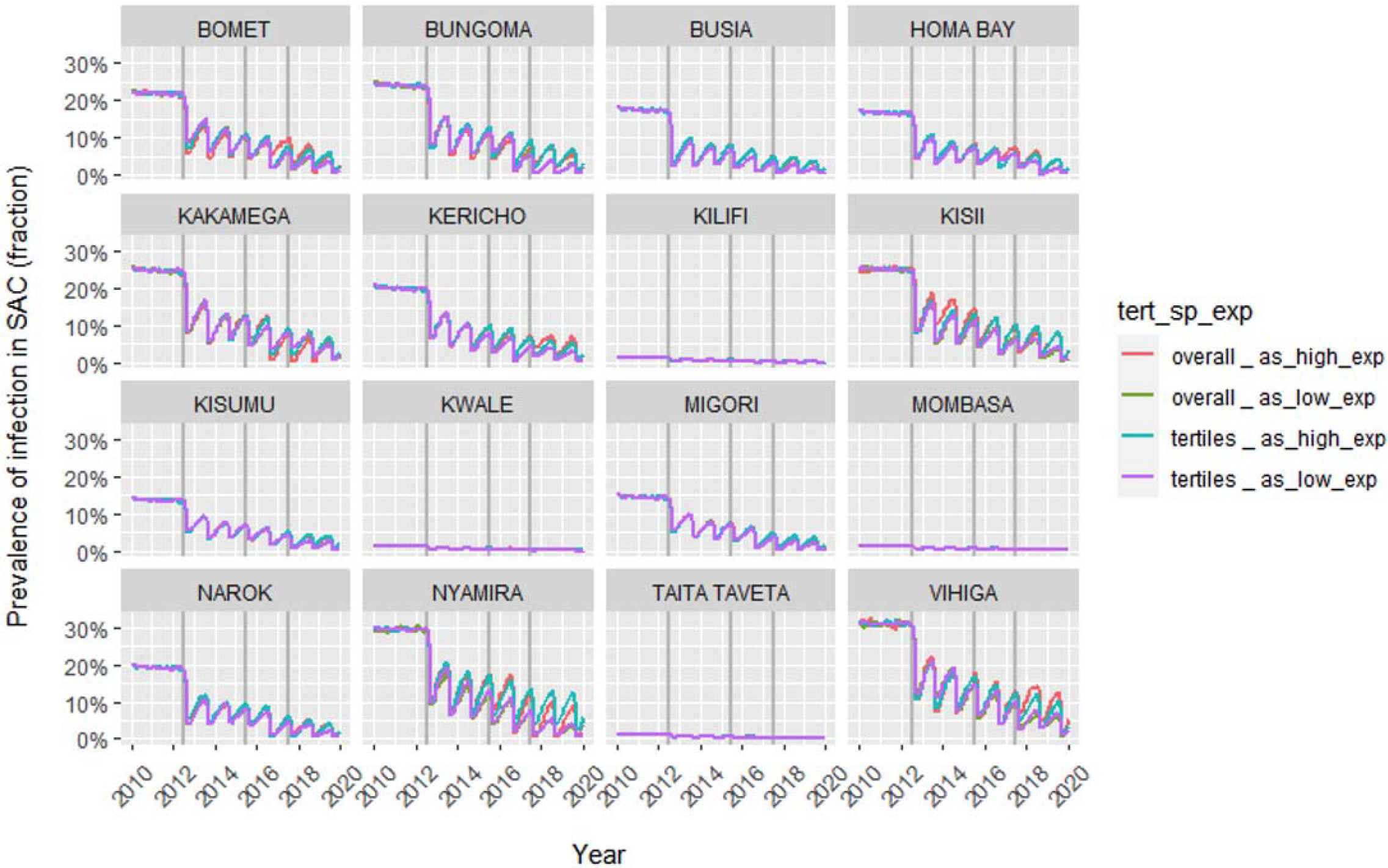
Predicted *A. lumbricoides* prevalence among school-aged children (SAC) at 16 IUs in Southwest Kenya over time, using 4 different modelling scenarios (described in Supplementary Table 2) with WORMSIM. Overall, the models modestly underestimated endline prevalence for the different UIs.

**Supplementary Figure 11.**
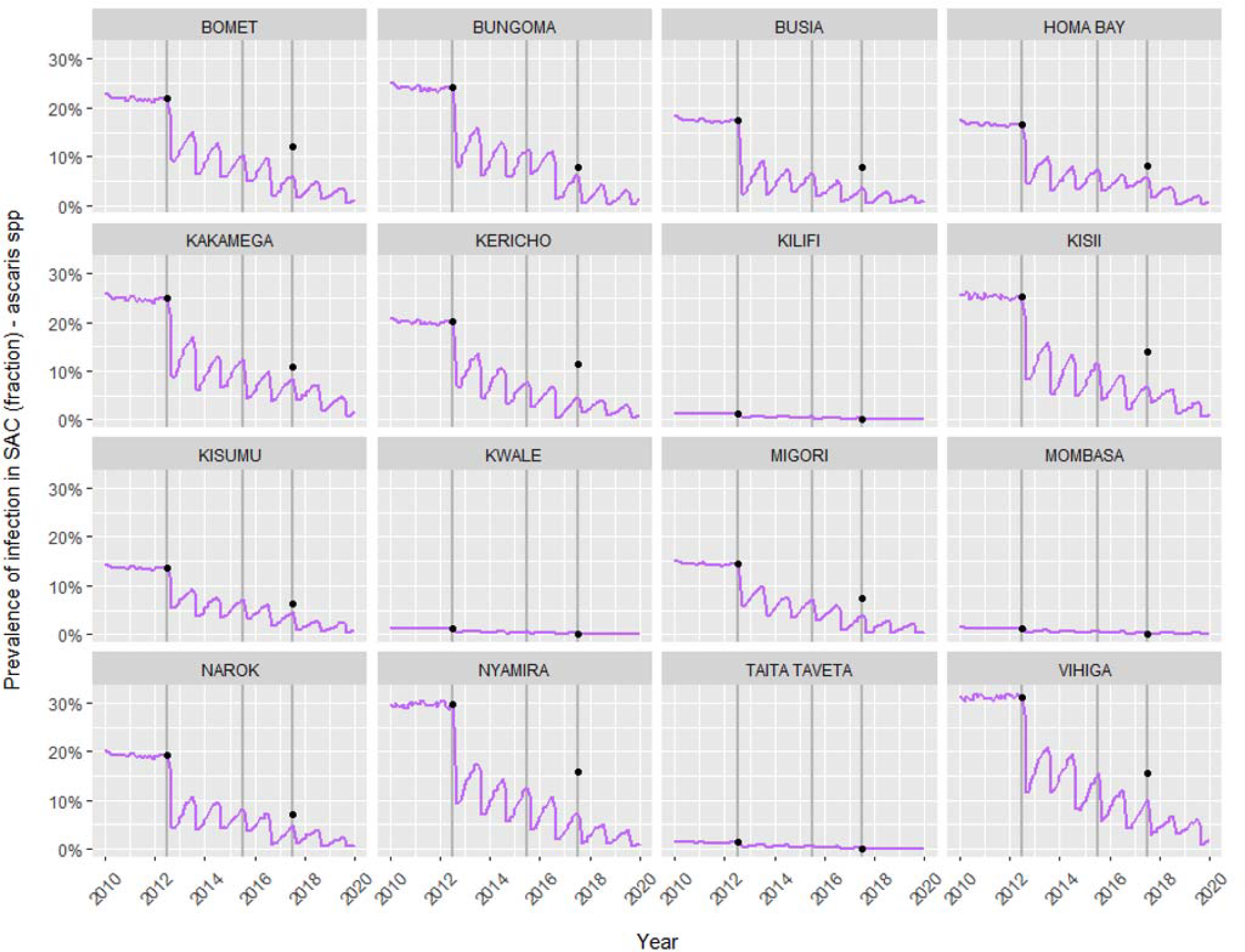
Predicted *A. lumbricoides* prevalence among school-aged children (SAC) at 16 IUs in Southwest Kenya over time, using WORMSIM. Black points show the measured prevalence levels among SAC at baseline and impact.

**Supplementary Figure 12.**
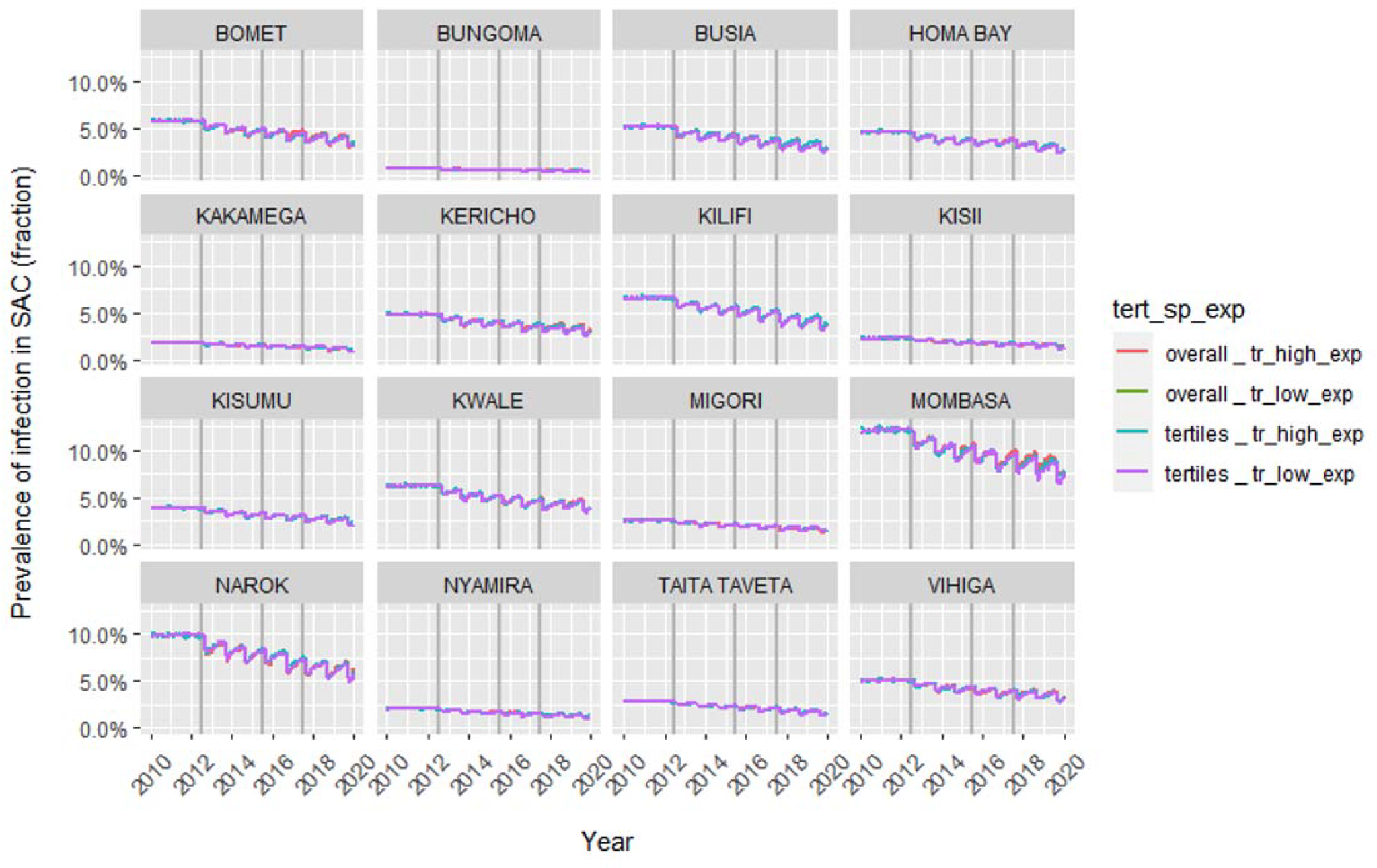
Predicted *T. Trichiura* prevalence among school-aged children (SAC) at 16 IUs in Southwest Kenya over time, using 4 different modelling scenarios (described in Supplementary Table 2) with WORMSIM.

For *T. trichiura*, in contrast to the predictions for *A. lumbricoides*, the model underestimated the prevalence reduction at impact in three IUs.

**Supplementary Figure 13.**
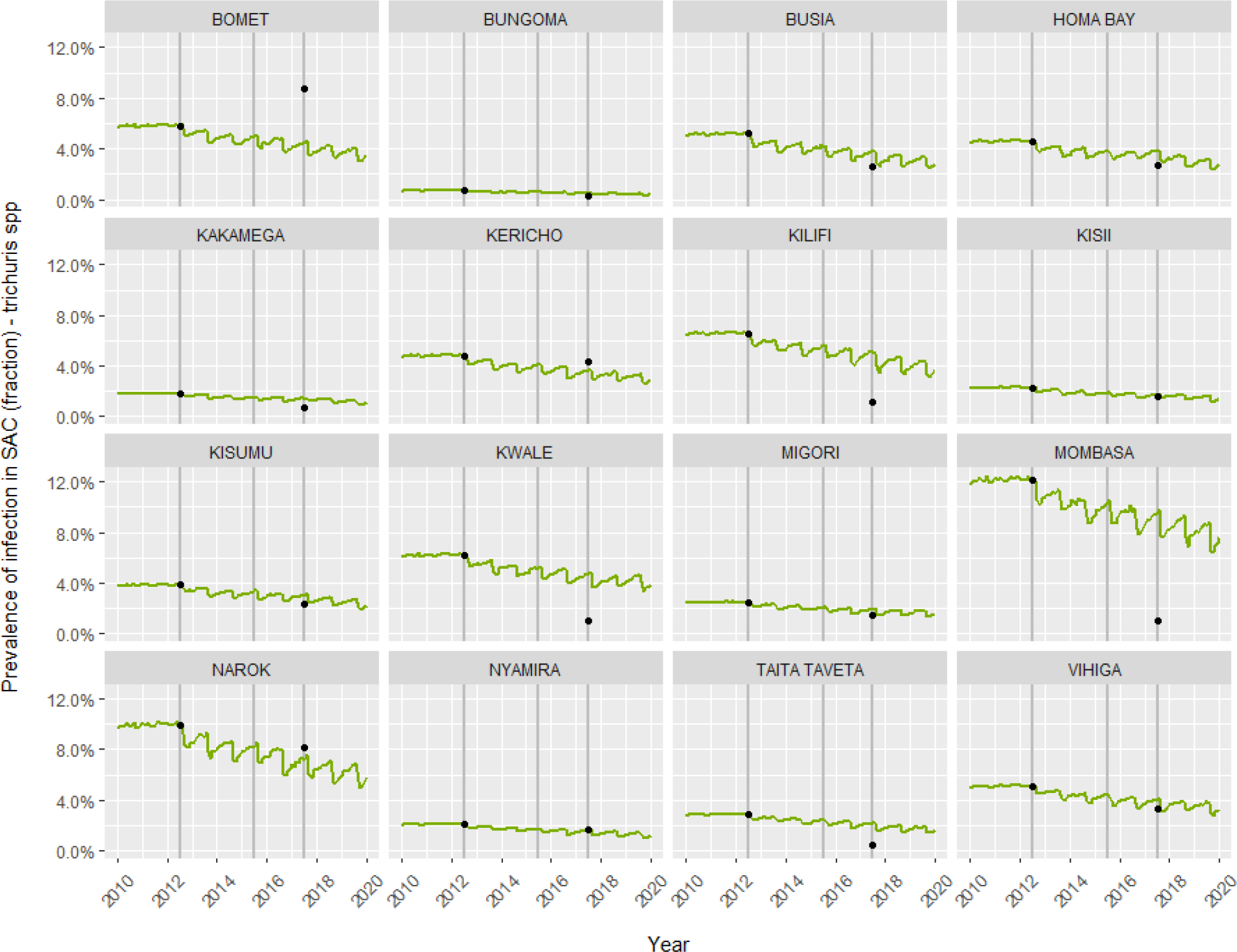
Predicted *T. trichiura* prevalence among school-aged children (SAC) at 16 IUs in Southwest Kenya over time, using WORMSIM. Black points show the measured prevalence levels among SAC at baseline and impact.

**Supplementary Figure 14.**
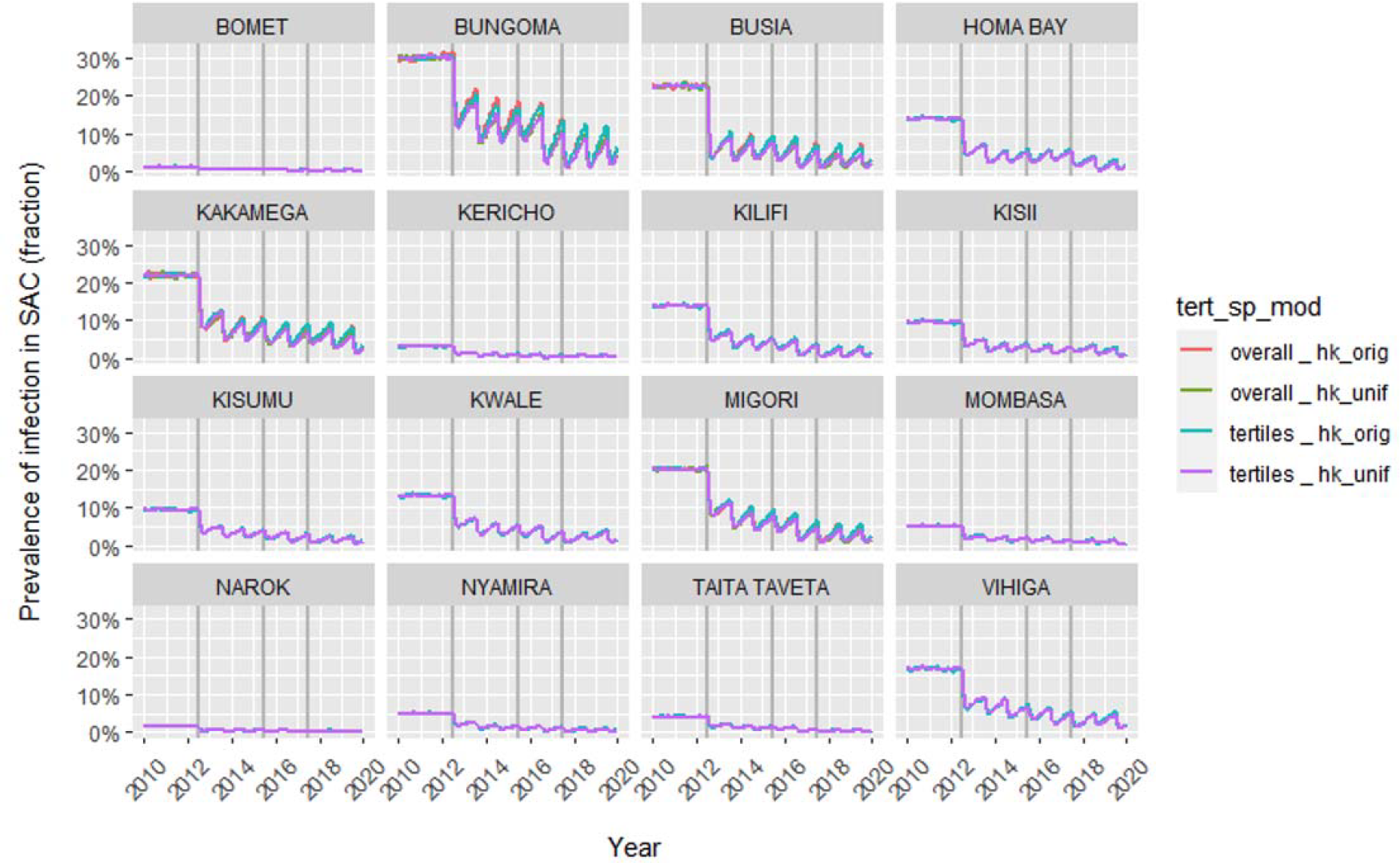
Predicted hookworm *spp*. prevalence among school-aged children (SAC) at 16 IUs in Southwest Kenya over time, using 4 different modelling scenarios (described in Supplementary Table 2) with WORMSIM.

There were no large differences in the predicted impact prevalence based on model #1 and model #3, the two scenarios assume different patterns of age-dependent exposure. Based on the predictions from model #1 (i.e Erasmus MC exposure function), prevalence in SAC is observed to go up faster after PC. This is due to larger exposure in adults compared to preSAC and SAC, creating a larger ‘pool’ of infected people in older people and hence higher infection pressure, resulting in quicker bounce-backs of prevalence after PC among SAC.

**Supplementary Figure 15.**
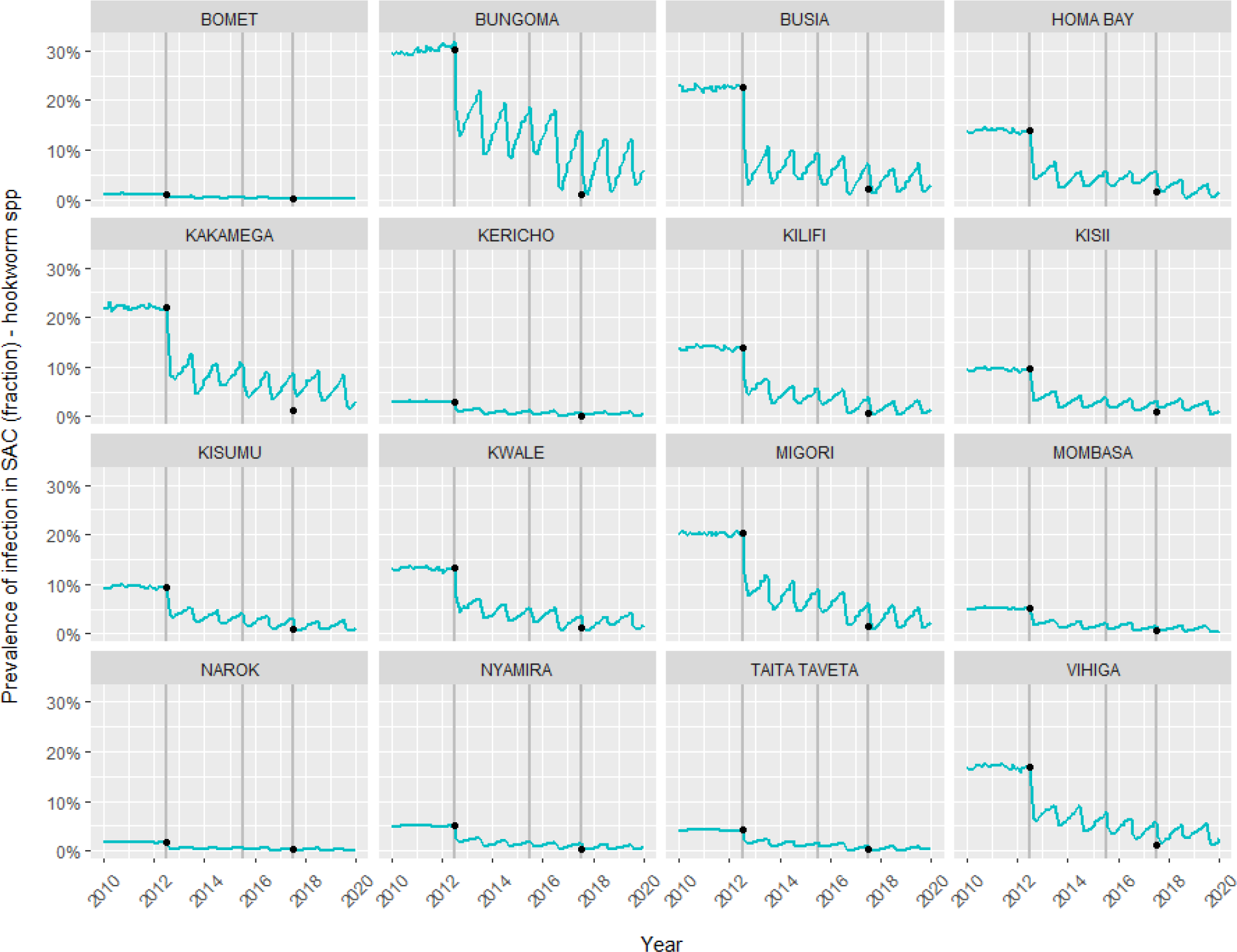
Predicted hookworm *spp*. prevalence among school-aged children (SAC) at the 16 IUs in Southwest Kenya over time, using WORMSIM. Black points show the measured prevalence levels among SAC at baseline and impact.

**Supplementary Figure 16.**
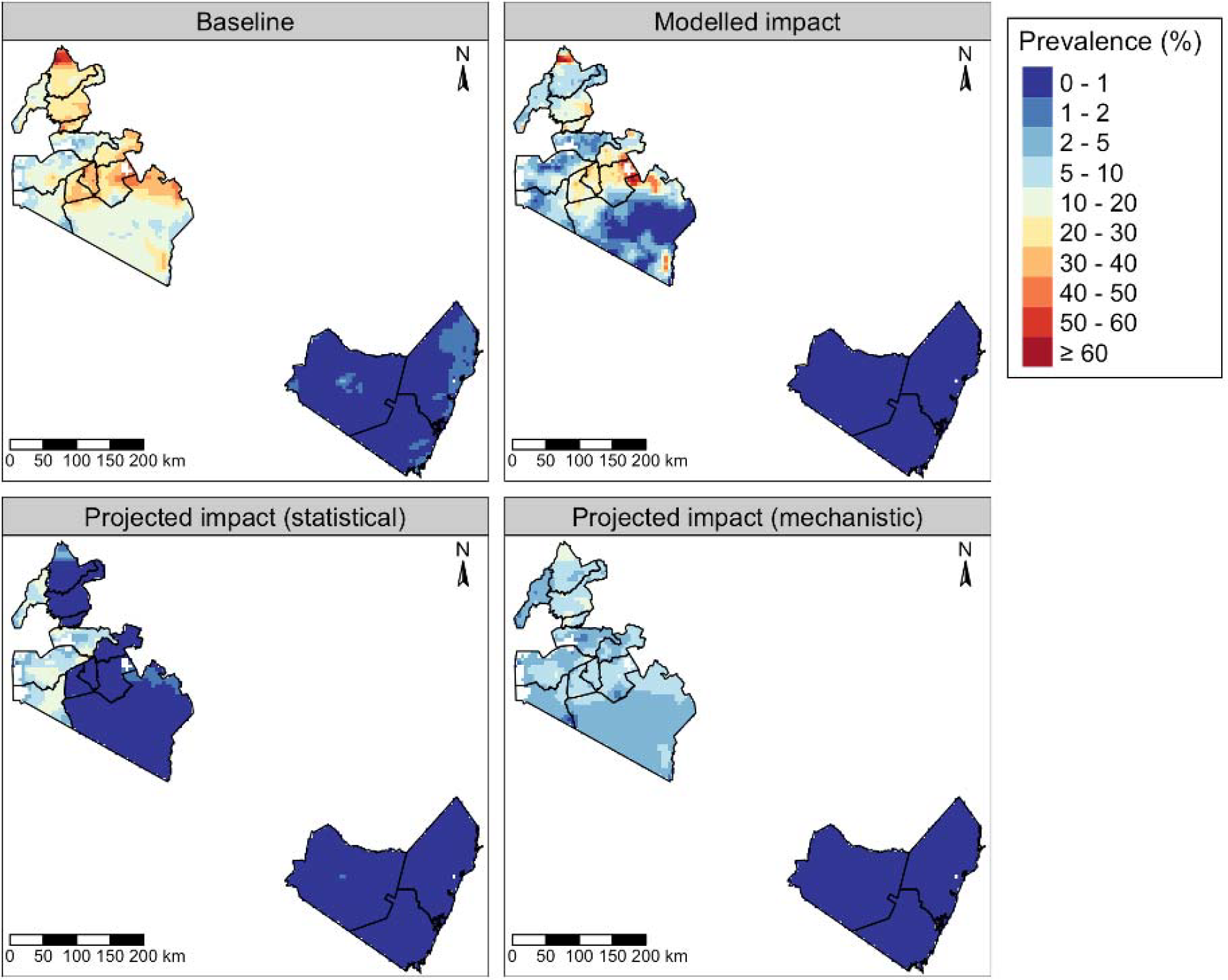
*A. lumbricoides* prevalence in SAC as predicted by the geostatistical model at baseline and impact, and as projected at impact using the statistical and mechanistic approaches.

**Supplementary Figure 17.**
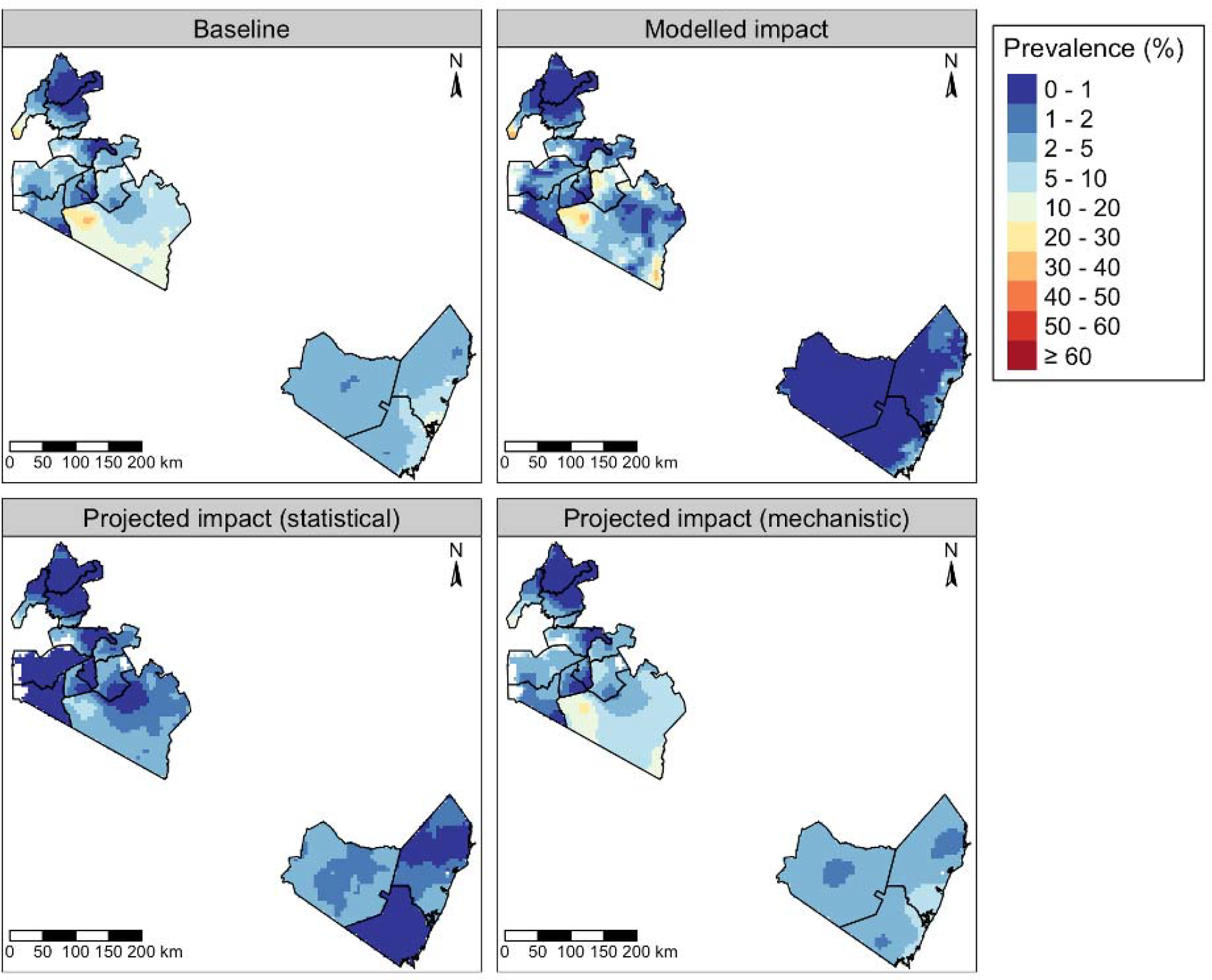
*T. trichiura* prevalence in SAC as predicted by the geostatistical model at baseline and impact, and as projected at impact using the statistical and mechanistic approaches.

**Supplementary Figure 18.**
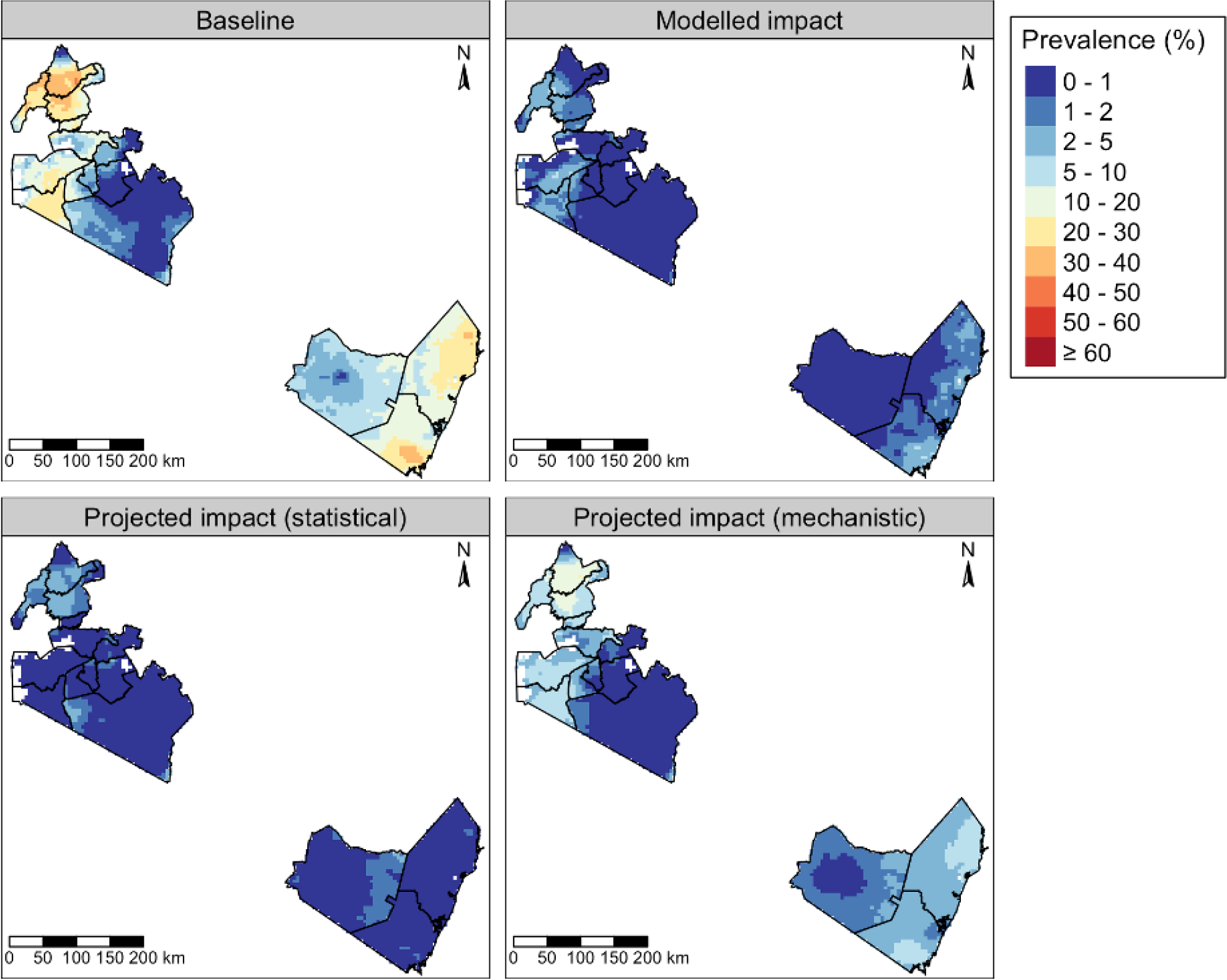
Hookworm *spp*. prevalence in SAC as predicted by the geostatistical model at baseline and impact, and as projected at impact using the statistical and mechanistic approaches.

